# Machine learning combining FIT with up to 1,025 clinical variables: limited referral reduction but potential for faster diagnosis

**DOI:** 10.64898/2026.07.16.26358105

**Authors:** Andres Tamm, Brian Shine, Tim James, Jaimie Withers, Hizni Salih, James E. East, Jason Oke, Jim Davies, Eva Morris, Brian D. Nicholson

## Abstract

**Background:** The faecal immunochemical test (FIT) is central to triaging symptomatic patients with suspected colorectal cancer (CRC) in UK primary care, yet only about one in eleven patients above the NICE 10 µg/g threshold have CRC. Existing prediction models attempting to improve on FIT have relied on conventional statistics and limited predictors.

**Methods:** GP-requested FITs with linked data (Jan 2017 - May 2025) were extracted from the Oxford University Hospitals (OUH) datawarehouse. Patients aged ≥18 with core bloods and 180-day CRC follow-up were included. Machine learning (ML) models were trained on up to 1,025 predictors: FIT, age, sex, blood tests and their time series slopes, diagnoses/procedures/prescriptions, deprivation, BMI, and ethnicity. Models comprised penalised logistic regression, generalised additive models (EBM, NAM, SNAM, NODE-GAM), decision tree ensembles (random forests, XGBoost), and a multilayer perceptron. Referral reduction versus FIT ≥10 µg/g was evaluated at model risk score thresholds capturing the same cancers (conservative) or same proportion of cancers (less conservative) as FIT. Potential to prioritise referred patients was assessed by examining whether positive predictive value (PPV) is very high (>30%) at any substantial sensitivity (>10%). Nested twice-repeated five-fold cross-validation provided unbiased estimates. An existing COLOFIT model was evaluated alongside.

**Findings:** 62,219 individuals (746 CRC) were analysed; 30,862 patients (315 CRC) with high/low risk symptoms and buffered FITs formed the primary subset. At ≥10 µg/g, FIT had 91.4% sensitivity, 84.2% specificity, 5.6% PPV, and 99.9% NPV. No model reduced referrals when required to capture the same cancers as in the FIT ≥10 µg/g cohort. Generalised additive models achieved up to 18.5% referral reduction when detecting the same proportion but some different cancers as FIT ≥10 µg/g (EBM: 18.5%, NODE-GAM: 17.5%, SNAM: 17.4%, COLOFIT: 16.7%). At 30% sensitivity, EBM, NAM and NODE-GAM had average PPVs between 34.6%-35.0%, while FIT had a PPV of 14.6%.

**Interpretation:** Generalised additive models (GAMs) reduced referrals on average by 19% if a small proportion of the FIT-positive CRCs were substituted with originally FIT-negative CRCs by the models. No model, including COLOFIT, reduced referrals while capturing all FIT-positive cancers. Generalised additive models could detect about a third of CRCs faster, as one in three patients flagged by the models had CRC at 30% sensitivity.

**Funding:** EPSRC Centre for Doctoral Training in Health Data Science; National Institute for Health Research (NIHR) Oxford Biomedical Research Centre; Cancer Research UK.

**Research in context:** *Evidence before this study:* The faecal immunochemical test (FIT) is widely used in primary care to triage symptomatic patients for colorectal cancer (CRC) investigations. Combining it with other routinely collected data (such age and blood tests) in prediction models may improve its performance. We used existing systematic reviews to understand the performance of FIT, and to identify CRC prediction models incorporating FIT in symptomatic primary care patients. We also searched PubMed and Google Scholar for machine learning models that incorporate FIT. We inferred that only about one in eleven symptomatic primary care patients have CRC, suggesting there is potential to improve precision and better target colonoscopy resources. Only three studies had developed models specifically in the target patient population, using traditional statistical methods with at most six predictor variables, indicating that machine learning methods and more diverse predictor variables remain unexplored in the target population.

*Added value of this study:* Using one of the largest samples to date (62,219 symptomatic primary care patients), we applied a range of interpretable and flexible machine learning models with up to 1,025 predictor variables (including blood test trends and medical history). We evaluated the models with clinically relevant performance metrics: potential to reduce colonoscopy referrals and potential to fast-track patients for investigations. No machine learning model or an existing COLOFIT model reduced referrals when required to detect all cancers already detected by FIT at the standard 10 µg/g threshold; the models reduced referrals at most by 19% if some missed FIT-positive cancers were substituted with extra FIT-negative cancers. However, when required to detect only 30% of cancers, generalised additive models had high positive predictive values (close to 35%), which were substantially higher than the PPV of FIT at the same sensitivity (14.6%). The models may thus be useful for fast-tracking higher-risk patients for colonoscopy to detect a subset of cancers faster.

*Implications of all the available evidence:* FIT is a highly sensitive and specific test for CRC: it is hard to improve on its performance with routinely collected data for reducing referrals in FIT-positive patients. However, prediction models show greater potential for prioritising referrals: detecting colorectal cancers faster in a subset of higher-risk patients.

## Background

The faecal immunochemical test (FIT) is recommended by the National Institute for Health and Care Excellence (NICE) for triaging patients with symptoms of colorectal cancer (CRC) in primary care for urgent cancer investigations^7^. These usually include colonoscopy, an invasive endoscopic examination of the lower gastrointestinal tract which has a small risk of serious complications. FIT measures the amount of haemoglobin (Hb) in stool, and results of at least 10 µg of Hb per gram are considered positive. However, despite its high sensitivity and specificity, only about 1 in 11 individuals who test positive have cancer (inferred from Booth et al data^1^). Colonoscopy services are under pressure, partly due to the after effects of the COVID-19 pandemic^8^. It is therefore worth investigating whether the positive predictive value (PPV) of FIT can be improved, to avoid unnecessary investigations and reduce pressure on endoscopy services. Prediction models that combine FIT with routinely collected data (such as age, sex, and blood tests) could potentially yield higher precision than FIT alone while capturing the same number of cancers. However, existing models developed in the target population^4–6^ (symptomatic patients with GP-requested FITs) have been limited to Cox and logistic models, and utilised at most six predictor variables.

We explore whether machine learning (ML) models can detect cancer better than FIT alone by (1) including diverse clinical predictor variables, (2) employing models with varying interpretability and flexibility, and (3) using novel ways of optimising the models. Diverse predictors—such as bloods, diagnosis and procedure codes—could leverage previously unconsidered relationships for more accurate CRC prediction. ML algorithms may perform better than logistic regression by capturing non-linear relationships (e.g. risk could be higher if blood test is both below or above reference range) and interactions (e.g. a test may be relevant only if another test is also abnormal). However, it is important to include interpretable ML models, to understand which variables drive the risk score. Finally, when a small proportion of patients have cancer, then optimising the models for ranking metrics (areas under the ROC and precision-recall curves) could outperform standard metrics, because ranking metrics are high when all individuals with cancer (no matter how few) have higher risk scores than individuals without cancer.

## Methods

### Ethics statement

The study was conducted as a service evaluation (OUH audit number 9076), and is reported following the TRIPOD-AI guidelines^9^.

### Data extraction

GP-requested quantitative FIT results (Jan 2017-May 2025) were extracted from the Oxford University Hospitals (OUH) Clinical Datawarehouse (CDW) and linked to secondary care and laboratory records. The resulting OUH-FIT dataset includes demographics, FIT, bloods, diagnoses, procedures, prescriptions, and histopathology reports. It covers nearly all GP FITs in Oxfordshire since the introduction of FIT in 2017, across approximately 76 general practices.

### Inclusion criteria

Eligible patients had a quantitative GP FIT, age ≥18 years, ≥180 days of follow-up from the earliest FIT, no prior CRC, and core bloods (haemoglobin [HGB], mean cell volume [MCV], platelets [PLT], white cells [WBC])^4^ within 180 days before earliest FIT. The 180-day follow-up condition was satisfied for all FITs received before 3/11/2024 (extraction date: 2/5/2025).

### Identification of colorectal cancers and clinical symptoms

CRC was identified using ICD-10 C18-C20 codes and pathology reports (rule-based algorithm^10^ with semi-manual verification as previously^11^). CRC treatments (radio-/chemotherapy, radical/local resection) were identified using OPCS-4 codes as previously^12^. The index date was the earliest of diagnosis-code, pathology, and treatment dates (for treatments ≤180 days before codes/reports). Symptoms prompting the FIT request were extracted from FIT request notes as previously^11^.

### Machine learning analysis

#### Outcome

Outcome was CRC detected within 180 days of the earliest FIT sample receipt, assuming most cancers present during FIT testing are detected by then given the NHS two-week wait and 28-day faster diagnosis standards. Time-to-event outcomes were not used as the aim was to infer if CRC was present at FIT testing rather than predict time to diagnosis; patients with short follow-up (e.g. 14 days) add little information to estimating 180-day CRC prevalence.

#### Predictors

Predictors were grouped into three sets (Table 1) to assess the value of incorporating more information:

1. FIT, age, sex and bloods available for ≥80% of patients (26 predictors; core bloods HGB, PLT, MCV, WBC available for all).
2. Set 1 plus blood test time series slopes over a period of 3 years pre-FIT available for ≥80% of patients (38 predictors).
3. Set 2 plus additional bloods and slopes within 1- and 3-year windows available for ≥20% of patients; clinical symptoms from FIT request notes; BMI, index of multiple deprivation (IMD), ethnicity; diagnosis (ICD-10), procedure (OPCS-4), and prescription codes recorded for ≥100 patients within prior 3 years (1,025 predictors).

**Table 1.**
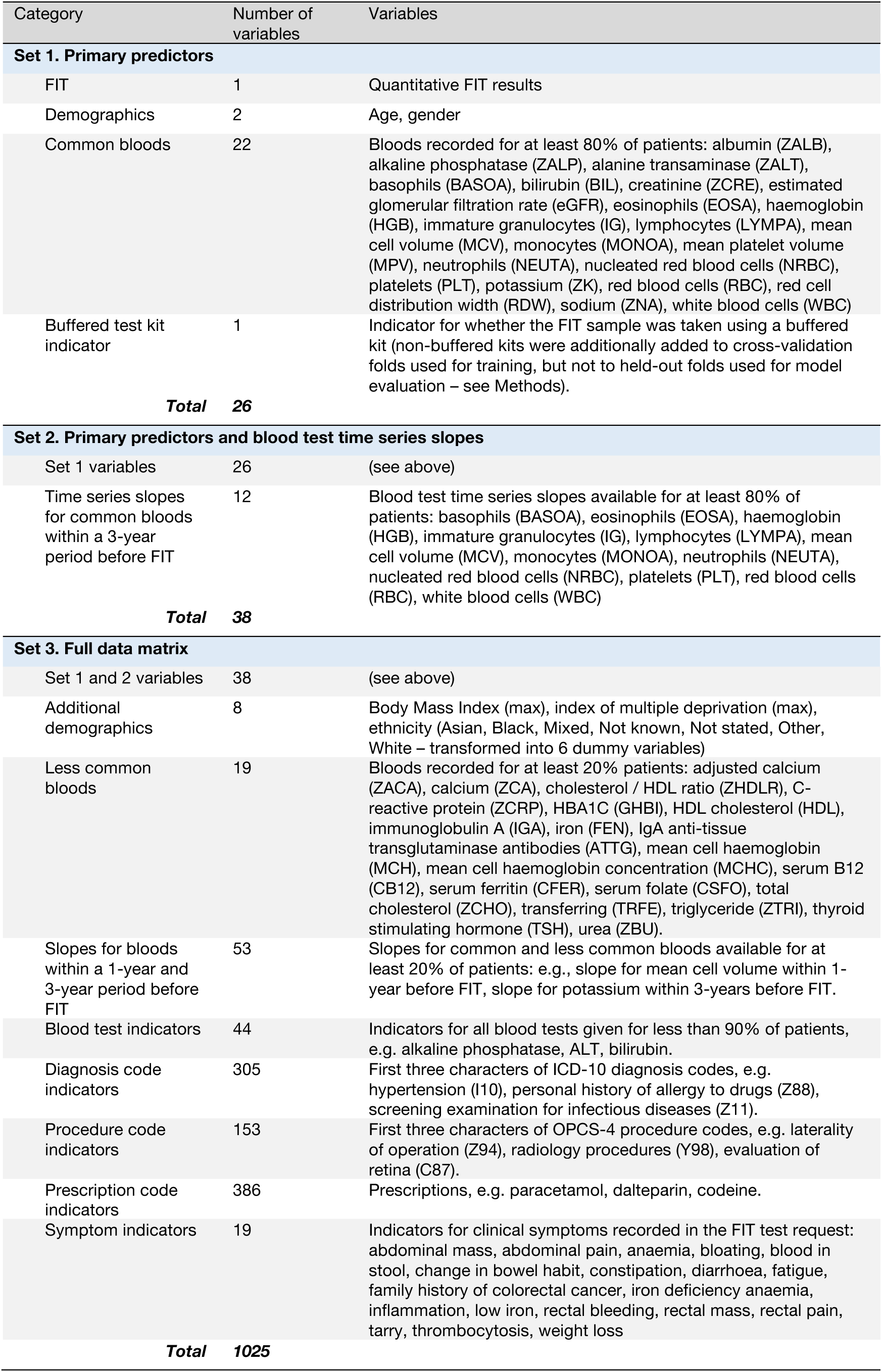
Predictor variables.

Models were trained on every set. For Set 3, models were trained on the full set and on 100 highest-ranked variables by XGBoost gain score, selecting the better configuration using validation (not test) scores to mitigate overfitting. If for any model Set 1 and Set 3 performances match, it implies the additional predictors do not add value.

#### Machine learning models

Models with varying flexibility and interpretability were included (Table 2): penalised logistic regression (PLR, elastic net)^13^, generalised additive models (GAMs), random forests (RF)^14^, gradient-boosted decision trees (XGBoost)^15^, and a multilayer perceptron (MLP). GAMs are fully interpretable yet more flexible than logistic regression and included the neural additive model (NAM)^16^, sparse neural additive model (SNAM)^17^, explainable boosting machine (EBM)^18^, and neural oblivious decision tree ensemble GAM (NODE-GAM)^19^. SNAM and NODE-GAM perform variable selection; NODE-GAM and EBM detect two-way interactions; and NAM, SNAM, and NODE-GAM can be optimised for ranking metrics (see below). RF and XGB were included for their strong performance across a range of datasets^20^. All models output a [0, 1]-interval risk score, interpretable as CRC probability if calibrated.

**Table 2.** Machine learning models.

| Model | Interpretability | Flexibility | Interactions |
| --- | --- | --- | --- |
| Linear models |  |  |  |
| Penalised logistic regression (PLR)<sup>13</sup> | Full | Low | None* |
| Generalised additive models (GAM) |  |  |  |
| Neural additive model (NAM)<sup>16</sup> | Full | Intermediate | None* |
| Sparse neural additive model (SNAM)<sup>17</sup> | Full | Intermediate | None* |
| Neural oblivious decision tree ensemble<br>GAM (NODE-GAM)<sup>19</sup> | Full | Intermediate | 2-way |
| Explainable boosting machine (EBM)<sup>18</sup> | Full | Intermediate | 2-way |
| Decision tree ensembles |  |  |  |
| Random forest (RF)<sup>14</sup> | Intermediate | High | 2-way and higher<br>order |
| Gradient-boosted decision tree (GBDT) -<br>xgboost<sup>15</sup> | Intermediate | High | 2-way and higher<br>order |
| Artificial neural networks |  |  |  |
| Multilayer perceptron (MLP) | Low | High | 2-way and higher<br>order |

Notes. \*Even though interaction terms can be included in logistic and neural additive models, this was not pursued in the data driven analysis because it is not clear which of the numerous possible variable combinations should be added. The other models allow detecting relevant interactions automatically.

#### Model selection and validation

Models were selected and validated using repeated five-fold cross-validation (CV) with an inner train-validation split for hyperparameter tuning (Figure 1). In each fold, 50 tuning trials were generated with tree-structured parzen estimators^21^. CV was repeated twice for robustness, but not more to reduce computation. Neural models were trained with a batch size of 1024 for up to 100 epochs with early stopping and exponential learning rate decay. Hyperparameters maximized validation set average precision (Supplement S1). Given low CRC prevalence, CV is more robust over a single split.

**Figure 1:**
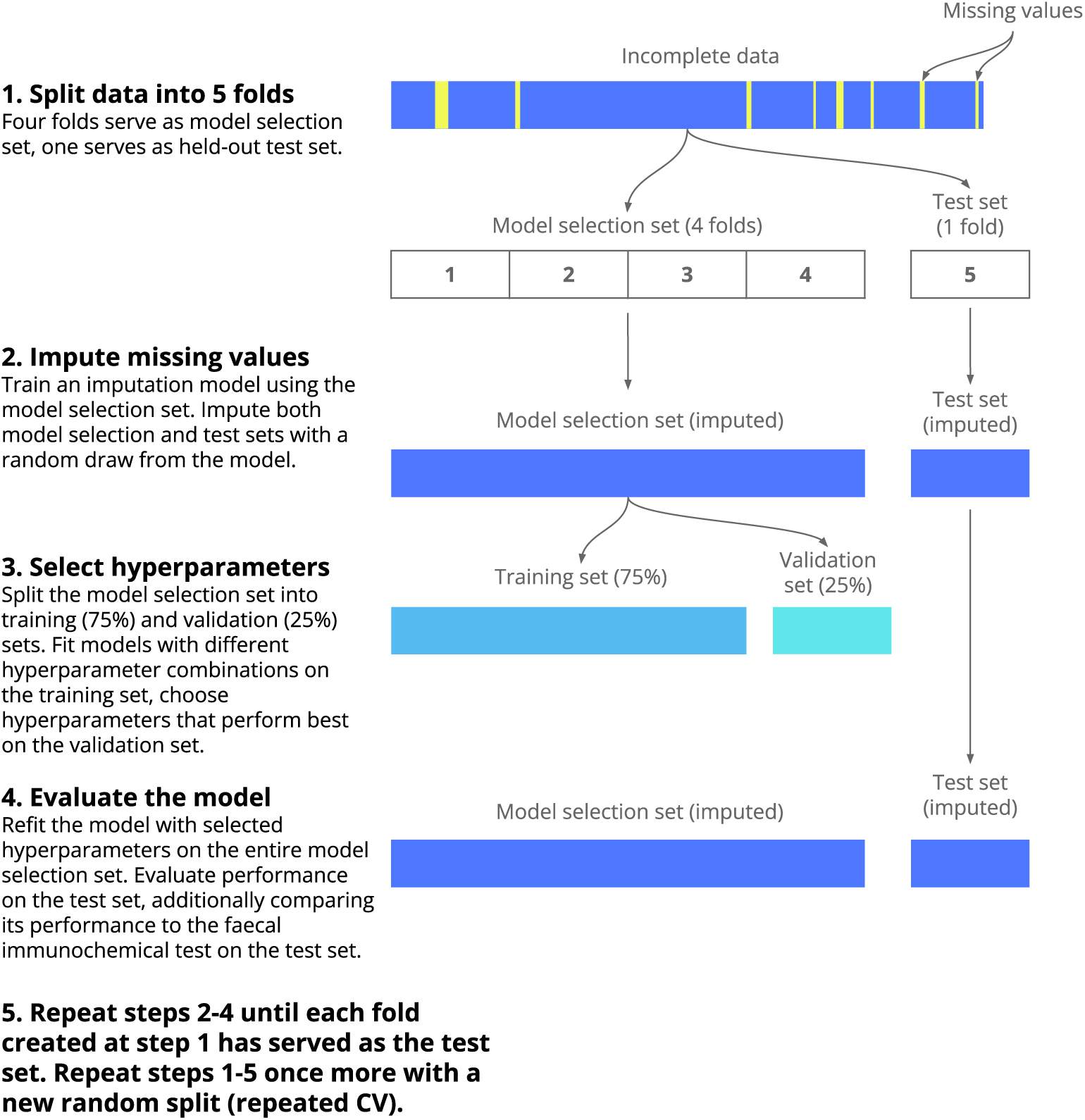
Model selection and evaluation using cross-validation with inner train-validation split. The data is first split into five folds; one serves as the held-out test set while the remaining serve as the model selection set. The model selection set is further split into training (75%) and validation sets (25%). The model is trained with different hyperparameter combinations on the training set; hyperparameters that perform best on the validation set are selected. The model with selected hyperparameters is then refitted on the entire model selection set and evaluated on the held-out test set. This outer cross-validation loop is repeated five times until each of the initial five folds has served as the held-out test set. The process is repeated once more (a different random split into five folds) for a more robust estimate of performance.

**Figure 2:**
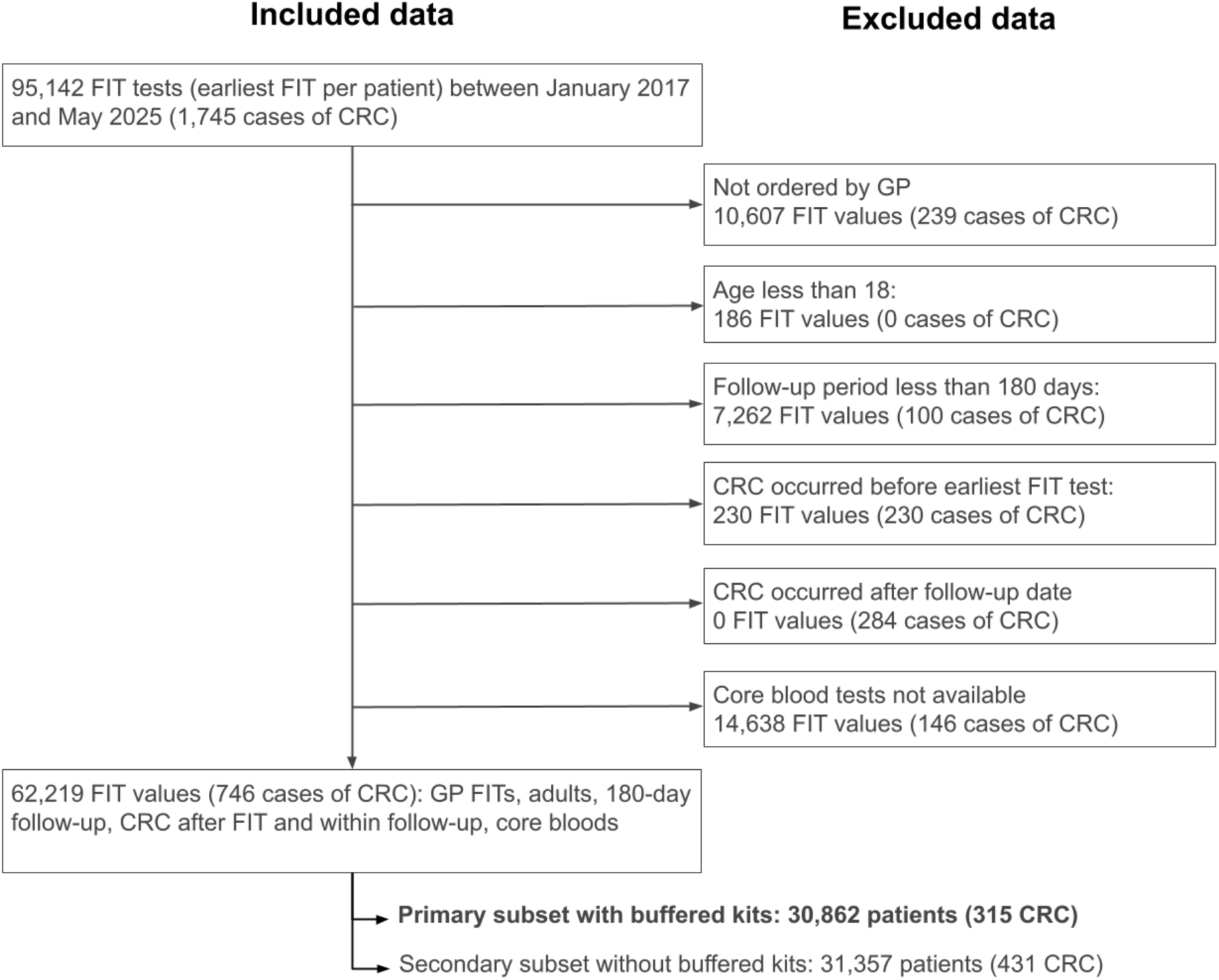
Number of patients and colorectal cancer cases at each step of applying the study inclusion criteria. The earliest FIT test result was used for each patient. CRC – colorectal cancer. Patients with CRC before FIT were excluded as they may already be at elevated risk compared to symptomatic patients without the history of CRC (target population). Cases of CRC that occurred after the 180-day follow-up were treated as non-CRCs in the model, because cases diagnosed with significant delay after FIT may be unrelated to the current symptoms, and treating these as CRCs would have meant that some patients are afforded longer follow-up than others which could bias estimates of CRC prevalence and model calibration.

#### Data preprocessing and transformations

Blood results were cleaned by removing date-like strings and non-numeric characters. Highly correlated bloods (r ≥ 0.9) were dropped. Tree-based models (EBM, RF, XGB) used raw predictors; others used transformations selected during tuning (log-transform plus standard scaling or quantile transform to Gaussian). All predictors were recorded at or before FIT sample receipt.

#### Clinically relevant model evaluation: referral reduction and referral prioritisation

FIT ≥10 µg/g achieves ≈90% sensitivity^1^ and is used to refer symptomatic patients for colorectal investigation. Models may improve upon FIT by capturing a similar number of cancers with less referrals. Referral reduction was assessed at model risk score thresholds capturing (1) the same cancers detected by FIT ≥10 µg/g (conservative), or (2) the same percentage of cancers as FIT ≥10 µg/g (less conservative, permitting substitution of FIT-positive cancers with FIT-negative cancers). At each threshold, the percentage reduction in positive tests relative to FIT was computed, reflecting potential decreases in downstream colonoscopy demand. The thresholds were estimated on held-out CV folds to equalise cancer detection across models (see Discussion). A test-reduction (TR) curve across the full sensitivity spectrum (0 – 100%) was also produced.

Models could also be used to prioritise referred FIT-positive patients for colorectal investigation, fast-tracking patients with higher risk scores to increase initial colonoscopy yield (without necessarily reducing referrals). To assess this, we examined whether each model achieved a high positive predictive value (PPV >30%) at thresholds capturing a substantial proportion of cancers (sensitivity >10%). We also visualised percent CRCs detected against percent patients investigated, when prioritising patients using model risk scores versus FIT values (Supplementary Materials S5). PPVs were obtained from precision-recall (PR) curves, with per-fold curves interpolated to a common sensitivity grid^22^.

#### Other performance metrics

Discrimination was assessed using the *c*-statistic, average precision (AP, area under the PR curve), ROC and PR curves. Calibration was summarised by observed/expected ratio, logistic slope and expected calibration error (ECE). An existing COLOFIT model that uses FIT, age, sex, platelets and mean cell volume as predictors was included for comparison^5^.

#### Optimisation for ranking metrics

For GAMs implemented in PyTorch^23^ (NAM, SNAM, NODE-GAM), novel methods were applied that optimise for high c-statistic and average precision^24,25^. This could yield better results when the proportion of cancers is small, because a traditional loss function can improve by giving non-cancer patients smaller risk scores even if they already rank below cancer patients.

#### Missing values

FIT, age, sex, and core bloods were complete by inclusion criteria, other predictors were partially missing. Within each CV fold, missing values were iteratively imputed using a MICE-like procedure with ridge regression on log-transformed predictors (Figure 1). Scikit-learn’s^26^ MICE was used to apply fitted imputers to held-out data; the method performed well in a recent study^27^. Data were imputed once per fold with different random draws. Cancer status was not used as it is unavailable at deployment.

#### Incorporating non-buffered FIT data

In Oxfordshire, FIT was offered for lower-risk symptoms, then extended to higher-risk symptoms from July 2020. Buffered kits preventing sample degradation were introduced in July 2021, test kit type was recorded from May 2022. To ensure that validation dataset reflects current practice (low- and high-risk [pan-risk] symptoms and buffered kits), the data with known buffered kits (since May 2022) were split into five cross-validation folds; older and non-buffered data were added into training folds only. Models included a buffered-kit indicator, and non-buffered records were optionally down-weighted during training (Supplement S2).

### Software and hardware

Analyses used Python 3.9 and relied on *pandas*^28^, *optuna*^21^, *interpret*^29^, *nodegam*^30^, PyTorch^23^, *scikit-learn*^26^, *xgboost*^15^, and *libauc*^31^. Code is available at https://github.com/tammandres/fitml. NAM, SNAM and NODE-GAM were trained on an NVIDIA A10 GPU.

## Findings

### Patient cohort

62,219 individuals (746 CRC) were included (Figure 1); 30,862 pan-risk patients (315 CRC) with buffered kits formed the primary subset. In the primary subset, cancer patients were older (CRC: median age 74.0, IQR 62.5.0-81.0; non-CRC: median 61.0, IQR 45.0-75.0), more often male (CRC: 57.5%, non-CRC: 39.7%), and more likely to have FIT ≥10 µg/g (CRC: 91.4%, non-CRC: 15.8%; Table 3). CRC patients were more likely to have anaemia, diarrhoea, rectal bleeding and blood in stool (Table 3). Individuals with CRC were also more likely to have low haemoglobin, elevated platelets, elevated white cells, low mean cell haemoglobin, low mean cell volume, low ferritin, and high C-reactive protein (Supplementary Table S3). In total, 5,128 (16.6%) patients in the primary subset had FIT ≥10 µg/g. At the threshold of ≥10 µg/g, FIT had 91.4% sensitivity, 84.2% specificity, 5.6% positive predictive value, and 99.9% negative predictive value. Descriptives for the entire patient cohort (primary and non-primary) are reported in Supplementary Table S4.

**Table 3.** Descriptive statistics for the patient cohort.

|  | No colorectal cancer | Colorectal cancer | Statistical test |
| --- | --- | --- | --- |
| Number of patients | 30547 | 315 | - |
| Age |  |  |  |
| 18-39.9 | 5054 (16.5%) | Not Available | - |
| 40-49.9 | 4410 (14.4%) | Not Available | - |
| 50-59.9 | 5153 (16.9%) | 42 (13.3%) | $\chi^2_{(1)}=2.8$, $p=0.095$ |
| 60-69.9 | 5127 (16.8%) | 53 (16.8%) | $\chi^2_{(1)}=0.0$, $p=0.984$ |
| 70-79.9 | 5938 (19.4%) | 101 (32.1%) | $\chi^2_{(1)}=31.6$, $p<0.001^{***}$ |
| $\geq 80$ | 4865 (15.9%) | 95 (30.2%) | $\chi^2_{(1)}=46.8$, $p<0.001^{***}$ |
| Median (25th, 75th) | 61.0 (45.0, 75.0) | 74.0 (62.5, 81.0) | - |
| Min, max | 18.0, 103.0 | 32.0, 94.0 | - |
| Gender |  |  |  |
| F (or not specified) | 18414 (60.3%) | 134 (42.5%) | $\chi^2_{(1)}=40.9$, $p<0.001^{***}$ |
| M | 12133 (39.7%) | 181 (57.5%) | $\chi^2_{(1)}=40.9$, $p<0.001^{***}$ |
| Ethnicity |  |  |  |
| Asian | 892 (2.9%) | Not Available | - |
| Black | 328 (1.1%) | Not Available | - |
| Mixed | 245 (0.8%) | - | - |
| Other Ethnic Groups | 366 (1.2%) | Not Available | - |
| White | 20981 (68.7%) | 214 (67.9%) | $\chi^2_{(1)}=0.1$, $p=0.776$ |
| Not stated | 6475 (21.2%) | 88 (27.9%) | $\chi^2_{(1)}=8.5$, $p=0.004^{**}$ |
| Not known | 1260 (4.1%) | Not Available | - |
| IMDD |  |  |  |
| Median (25th, 75th) | 8.0 (6.0, 10.0) | 8.0 (7.0, 10.0) | - |
| Min, max | 1.0, 10.0 | 1.0, 10.0 | - |
| Not known | 5519 (18.1%) | 24 (7.6%) | $\chi^2_{(1)}=23.1$, $p<0.001^{***}$ |
| FIT ($\mu\text{g Hb/g}$) | | | |
| 0-1.9 | 20959 (68.6%) | 15 (4.8%) | $\chi^2_{(1)}=583.8$, $p<0.001^{***}$ |
| 2-9.9 | 4748 (15.5%) | 12 (3.8%) | $\chi^2_{(1)}=32.9$, $p<0.001^{***}$ |
| 10-99.9 | 3237 (10.6%) | 93 (29.5%) | $\chi^2_{(1)}=116.0$, $p<0.001^{***}$ |
| $\geq 100$ | 1603 (5.2%) | 195 (61.9%) | $\chi^2_{(1)}=1824.2$, $p<0.001^{***}$ |
| Median (25th, 75th) | 0.0 (0.0, 3.0) | 304.0 (47.2, 400.0) | - |
| Min, max | 0.0, 400.0 | 0.0, 400.0 | - |
| Symptoms - GP reported |  |  |  |
| Abdominal mass | 43 (0.1%) | - | - |
| Abdominal pain | 3784 (12.4%) | 29 (9.2%) | $\chi^2_{(1)}=2.9$, $p=0.088$ |
| Anaemia | 4072 (13.3%) | 57 (18.1%) | $\chi^2_{(1)}=6.1$, $p=0.013^*$ |
| Bloating | 1077 (3.5%) | Not Available | - |
| Blood in stool | 4211 (13.8%) | 70 (22.2%) | $\chi^2_{(1)}=18.6$, $p<0.001^{***}$ |
| Change in bowel habit | 6709 (22.0%) | 70 (22.2%) | $\chi^2_{(1)}=0.0$, $p=0.912$ |
| Constipation | 986 (3.2%) | Not Available | - |
| Diarrhoea | 2977 (9.7%) | 20 (6.3%) | $\chi^2_{(1)}=4.1$, $p=0.043^*$ |
| Family history of CRC | 244 (0.8%) | - | - |
| Fatigue | 682 (2.2%) | Not Available | - |
| Inflammation | 361 (1.2%) | Not Available | - |
| Iron deficiency anaemia | 2038 (6.7%) | 31 (9.8%) | $\chi^2_{(1)}=5.0$, $p=0.025^*$ |
| Low iron | 1347 (4.4%) | 15 (4.8%) | $\chi^2_{(1)}=0.1$, $p=0.762$ |
| Melaena | 246 (0.8%) | Not Available | - |
| Rectal bleeding | 2741 (9.0%) | 48 (15.2%) | $\chi^2_{(1)}=14.9$, $p<0.001^{***}$ |
| Rectal mass | 20 (0.1%) | Not Available | - |
| Rectal pain | 159 (0.5%) | Not Available | - |
| Thrombocytosis | 362 (1.2%) | Not Available | - |
| Weight loss | 2352 (7.7%) | 20 (6.3%) | $\chi^2_{(1)}=0.8$, $p=0.371$ |
| Not known | 4665 (15.3%) | 41 (13.0%) | $\chi^2_{(1)}=1.2$, $p=0.268$ |
| CRC-relevant treatments* |  |  |  |
| No treatments | 26857 (87.9%) | 42 (13.3%) | $\chi^2_{(1)}=1549.8$, $p<0.001^{***}$ |
| Chemotherapy | 887 (2.9%) | 145 (46.0%) | $\chi^2_{(1)}=1794.3$, $p<0.001^{***}$ |
| Local excision | 32 (0.1%) | 15 (4.8%) | $\chi^2_{(1)}=444.7$, $p<0.001^{***}$ |
| Radical resection | 115 (0.4%) | 181 (57.5%) | $\chi^2_{(1)}=10695.5$, $p<0.001^{***}$ |
| Radiotherapy | 665 (2.2%) | 60 (19.0%) | $\chi^2_{(1)}=386.8$, $p<0.001^{***}$ |
| T-stage (extracted)** |  |  |  |
| 0 | - | Not Available | - |
| 1 | - | 34 (10.8%) | - |
| 2 | - | Not Available | - |
| 3 | - | 99 (31.4%) | - |
| 4 | - | 40 (12.7%) | - |
| Not known | - | 108 (34.3%) | - |

Notes. \*CRC-relevant treatments are procedures used for treating colorectal cancer (CRC), but they may also be given for other conditions. \*\*T-stage was extracted from radiology and pathology reports using a pattern-matching algorithm. "Not available" means that the count was $< 10$ , in which case the category with the second lowest count was hidden too, so that it could not be derived from the total; to comply with OUH requirements.

### Potential to reduce referrals

Reduction in referrals was first evaluated at equivalent sensitivity: can the models detect the same percentage of cancers as FIT ≥10 µg/g, while generating fewer referrals? (Some FIT-positive cancers may be missed if capturing extra FIT-negative cancers.) EBM yielded the highest reduction (-18.5%, standard deviation [SD] across the ten folds = 11.8%), followed by NODE-GAM (-17.5%, SD = 18.6%), SNAM (-17.4%, SD = 11.4%), NAM (-15.5%, SD = 16.1%), RF (-13.9%, SD = 12.7%), PLR (-6.6%, SD = 23.5%), and XGB (-3.5%, SD = 20.9%) (Figure 3: top panel, Table 4). MLP increased referrals (11.1, SD = 21.7%). COLOFIT yielded a -16.7% reduction (SD = 9.1%). COLOFIT missed a small proportion of FIT-positive CRCs in 7 out of 10 folds, EBM and NODE-GAM in 10 folds; this constituted fewer than 10 (<18%) CRCs (data cannot be reported for fewer than ten patients per OUH requirements). Some missed FIT-positive CRCs and some detected FIT-negative CRCs were locally advanced (pathological T-stage 3-4). Precise comparison requires larger sample and more complete TNM-staging data.

**Figure 3.**
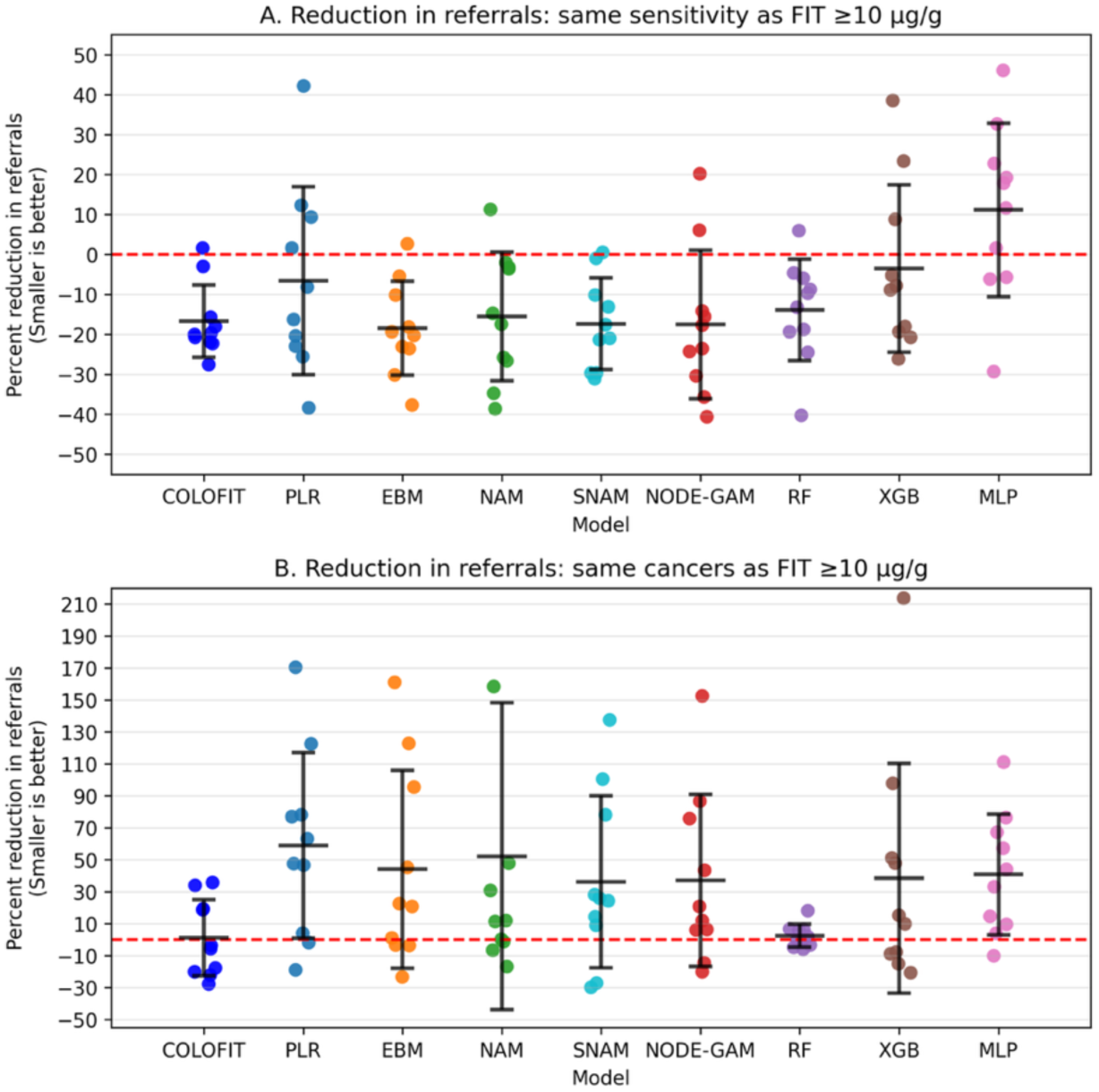
Potential reduction in referrals for the machine learning models and COLOFIT. Reduction in referrals is the percent reduction in patients testing positive relative to FIT ≥10 µg/g, at model risk-score thresholds chosen to capture a comparable number of cancers. **A:** Reduction in referrals at thresholds that yield the same sensitivity as FIT ≥10 µg/g, so a model may miss some FIT-positive cancers if it captures extra FIT-negative cancers (less conservative). **B**: Reduction in referrals at thresholds that capture the same cancers as FIT ≥10 µg/g, so a model is required to detect all cancers already detected by FIT ≥10 µg/g (more conservative). Values below zero indicate fewer referrals while capturing a comparable number of cancers; values above zero indicate more referrals (undesirable). Points show the ten cross-validation folds; the central horizontal line is the mean and whiskers span ±1 standard deviation across folds. Abbreviations: FIT – faecal immunochemical test, COLOFIT – an existing prediction model, PLR – penalised logistic regression, EBM – explainable boosting machine, NAM – neural additive model, SNAM – sparse neural additive model, RF – random forests, XGB – XGBoost, MLP – multilayer perceptron.

**Table 4.** Reduction in referrals.

| Model name | Reduction in referrals relative to FIT $\geq 10$ $\mu\text{g/g}$ (same sensitivity); smaller values mean higher reduction | Reduction in referrals relative to FIT $\geq 10$ $\mu\text{g/g}$ (same cancers); smaller values mean higher reduction |
| --- | --- | --- |
| COLOFIT | -16.7 (9.1) | 1.2 (23.9) |
| PLR | -6.6 (23.5) | 58.9 (58.0) |
| **EBM** | **-18.5 (11.8)** | 44.0 (61.9) |
| NAM | -15.5 (16.1) | 52.2 (96.0) |
| SNAM | -17.4 (11.4) | 36.2 (53.7) |
| NODE-GAM | -17.5 (18.6) | 37.0 (53.8) |
| RF | -13.9 (12.7) | 2.5 (7.2) |
| XGB | -3.5 (20.9) | 38.4 (71.9) |
| MLP | 11.1 (21.7) | 40.8 (37.7) |

Notes. Reduction in referrals (same sensitivity) shows the percent reduction in the number of patients testing positive at a model risk score threshold that captures the same proportion of cancers as FIT $\geq 10$ $\mu\text{g/g}$ (equal sensitivity). Reduction in referrals (same cancers) shows the percent reduction at a threshold that captures the same cancers as FIT $\geq 10$ $\mu\text{g/g}$ . Each cell shows mean and standard deviation (in brackets) across the ten cross-validation folds. Highest average reduction is shown in bold (no models produced reduction on average when required to detect all FIT-positive cancers). Models: COLOFIT - an existing prediction model, PLR - penalised logistic regression, EBM - explainable boosting classifier, NAM - neural additive model, SNAM - sparse neural additive model, NODE-GAM - neural oblivious decision tree ensemble GAM, RF - random forests, XGB - XGBoost, MLP - multilayer perceptron.

Secondly, we evaluated referral reduction conservatively: can the models detect cancers identified by FIT ≥10 µg/g while reducing referrals. No model, including COLOFIT, achieved a reduction in referrals across the ten cross-validation folds on average (Figure 3: bottom panel; Table 4: ‘Reduction in referrals (same cancers)’). This indicates that the models were unable to detect all individuals with a positive FIT who had cancer while reducing referrals.

### Potential to prioritise referred patients

Potential to prioritise referred patients is approximately indicated by high PPVs (>30%) at sensitivities where more than 10% of cancers are detected. For example, at 10% sensitivity, EBM, NAM and NODE-GAM all had average PPVs close to 50%: EBM (48.3%, SD = 14.2%), NAM (49.9%, SD = 16.4%), and NODE-GAM (49.4%, SD = 20.4%), while FIT had a PPV of 14.6% (SD = 1.5%). Similarly, at 30% sensitivity, EBM, NAM and NODE-GAM had average PPVs close to 35%: EBM (34.6%, SD = 6.5%), NAM (34.7%, SD = 6.1%), NODE-GAM (35.0%, SD = 8.4%), while FIT again had a PPV of 14.6% (SD = 1.5%). See Table 5 and Figure 4 for the PPV-sensitivity trade-off. EBM, NAM and NODE-GAM may thus be effective for detecting at least a third of CRCs faster, as on average, at least one in three flagged by the models had CRC at 30% sensitivity (Table 5; Figure 4: Panel B). A simplified simulation showed that prioritising patients using NODE-GAM risk-scores led to faster initial CRC detection compared to FIT values (Supplementary Materials S5).

**Figure 4.**
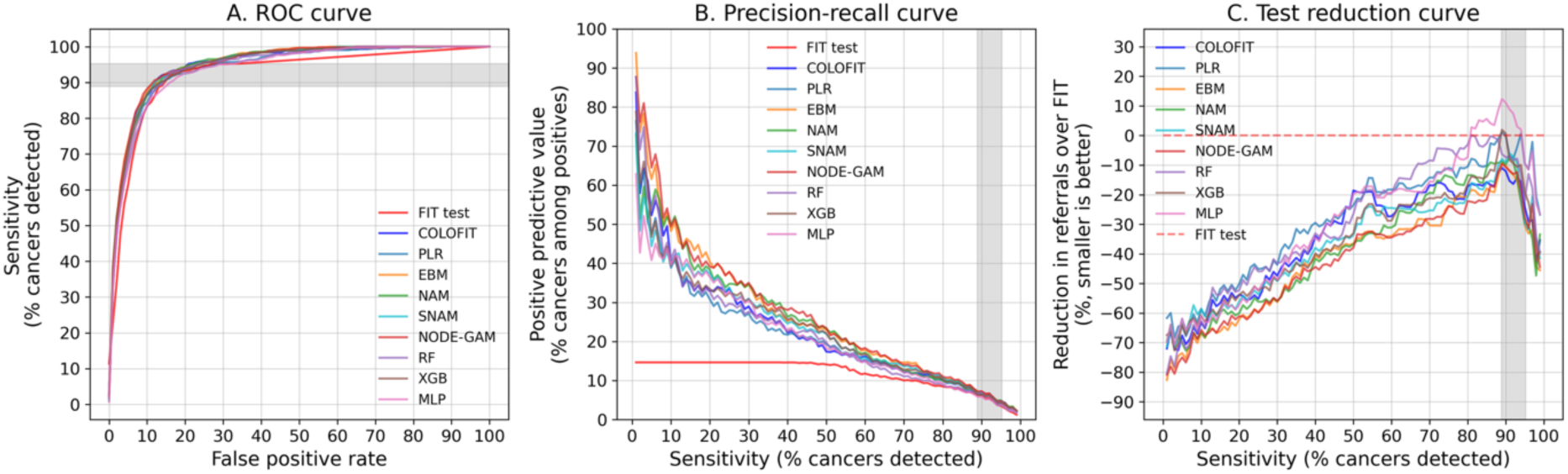
Receiver-operating characteristic (ROC), precision-recall and test-reduction curves for machine learning models, COLOFIT and the FIT test. Curves show the mean value of each quantity over ten held-out cross-validation folds. The vertical grey area is shaded between the minimum and maximum sensitivities of FIT ≥10 µg/g in the held-out folds. The test reduction curve shows the percent reduction in the number of positive tests relative to FIT at each level of sensitivity (the model and FIT thresholds yield the same sensitivity). All curves were interpolated to a fixed grid of values at 1-unit increments. Abbreviations: FIT – faecal immunochemical test, COLOFIT – an existing prediction model, PLR – penalised logistic regression, EBM – explainable boosting machine, NAM – neural additive model, SNAM – sparse neural additive model, RF – random forests, XGB – XGBoost, MLP – multilayer perceptron. Standard deviation across the cross-validation folds is not shown here to make the figure easier to inspect – please see Supplementary Figure S6 that includes average plus-minus one standard deviation.

**Table 5.** Positive predictive value at sensitivities < 50%.

| Model | PPV-10 | PPV-20 | PPV-30 | PPV-40 | PPV-50 |
| --- | --- | --- | --- | --- | --- |
| FIT test | 14.6 (1.5) | 14.6 (1.5) | 14.6 (1.5) | 14.6 (1.5) | 14.1 (1.8) |
| COLOFIT | 39.8 (2.8) | 33.0 (6.7) | 29.0 (6.4) | 22.3 (2.6) | 17.4 (2.1) |
| PLR | 38.9 (12.5) | 30.8 (6.6) | 27.2 (5.7) | 21.7 (4.8) | 19.3 (4.0) |
| **EBM** | 48.3 (14.2) | **40.4 (9.0)** | 34.6 (6.5) | 25.2 (4.6) | 23.0 (4.4) |
| **NAM** | **49.9 (16.4)** | 38.4 (8.0) | 34.7 (6.1) | 26.3 (5.9) | 23.3 (4.9) |
| SNAM | 39.9 (14.2) | 37.4 (12.1) | 31.0 (7.4) | 24.6 (5.2) | 21.9 (4.5) |
| **NODE-GAM** | 49.4 (20.4) | 39.1 (7.9) | **35.0 (8.4)** | **28.1 (7.7)** | **23.4 (4.2)** |
| RF | 41.3 (8.3) | 32.8 (8.7) | 27.6 (7.5) | 22.5 (4.5) | 19.7 (4.0) |
| XGB | 39.5 (9.7) | 33.4 (8.1) | 30.8 (9.2) | 25.6 (6.3) | 22.1 (5.4) |
| MLP | 39.7 (10.5) | 36.5 (10.7) | 30.5 (8.7) | 23.0 (4.4) | 18.2 (3.8) |

Notes. PPV-10 - Positive predictive value (PPV) at 10% sensitivity; PPV-20 - PPV at 20% sensitivity. Each cell shows mean and standard deviation (in brackets) across the ten cross-validation folds. Highest average scores in each column are highlighted in bold. Models: COLOFIT - an existing prediction model, PLR - penalised logistic regression, EBM - explainable boosting classifier, NAM - neural additive model, SNAM - sparse neural additive model, NODE-GAM - neural oblivious decision tree ensemble GAM, RF - random forests, XGB - XGBoost, MLP - multilayer perceptron.

### Discrimination

Average precision (AP) summarises area under the precision-recall (PR) curve and is suitable for low-prevalence data^22^. NODE-GAM achieved the highest AP of 27.6% (SD = 3.8%), followed by EBM (27.0%, SD = 3.3%) and NAM (25.7%, SD = 4.6%). APs for other models were lower (Table 6); COLOFIT yielded an AP of 23.5% (SD = 2.7%); FIT had an AP of 11.8% (SD = 1.2%). The highest c-statistics were obtained by EBM (95.1%, SD = 0.6%), NODE-GAM (95.0%, SD = 0.6%), and NAM (94.8%, SD = 0.4%) – see Table 6 for other models. COLOFIT achieved a c-statistic of 94.7% (SD = 0.8%); FIT 92.2% (SD = 1.2%). The models and FIT were not well-distinguished visually on the ROC curves (Figure 4: Panel A). The PR curves (Figure 4: Panel B) revealed that all models had higher average PPVs than FIT at sensitivities < 50%. ROC and PR-curves showing variability across the cross-validation folds can be found in Supplementary Figure S6.

**Table 6.** Discrimination metrics.

| Model name | Average precision | c-statistic |
| --- | --- | --- |
| FIT test | 11.8 (1.2) | 92.2 (1.2) |
| COLOFIT | 23.5 (2.7) | 94.7 (0.8) |
| PLR | 22.1 (4.3) | 93.9 (1.1) |
| **EBM** | 27.0 (3.3) | **95.1 (0.6)** |
| NAM | 25.7 (4.6) | 94.8 (0.4) |
| SNAM | 24.0 (4.2) | 94.8 (0.7) |
| **NODE-GAM** | **27.6 (3.8)** | 95.0 (0.6) |
| RF | 23.1 (2.9) | 94.1 (1.0) |
| XGB | 24.0 (3.8) | 94.6 (1.0) |
| MLP | 22.5 (3.7) | 93.7 (0.7) |

*Notes.* Each cell shows mean and standard deviation (in brackets) across the ten cross-validation folds. Highest average scores in each column are highlighted in bold. Models: COLOFIT - an existing prediction model, PLR - penalised logistic regression, EBM - explainable boosting classifier, NAM - neural additive model, SNAM - sparse neural additive model, NODE-GAM - neural oblivious decision tree ensemble GAM, RF - random forests, XGB - XGBoost, MLP - multilayer perceptron.

### Most informative predictor variables

In the best performing models (EBM, NODE-GAM), the ten most important variables across the cross-validation folds were FIT, age, sex, change in bowel habit (yes/no), changes in MCV and mean cell haemoglobin (MCH) over a 3-year period before FIT, PLT, albumin, serum ferritin, and the urea blood test order (yes/no). The association between these variables and cancer risk is visualised in Supplementary Materials S7.

### Sensitivity analyses and additional results

Of the highest performing models (EBM, NODE-GAM), EBM was well calibrated on average, while NODE-GAM underpredicted risk (Supplementary Table S8). Fine-tuning NODE-GAM for high areas under the PR and ROC curves did not yield better referral reduction or prioritisation (Supplementary Figure S9). The inclusion of additional predictors (Set 1 -> Set 3) increased the referral prioritisation potential of EBM, leading to higher PPVs at <50% sensitivity (Supplementary Figure S10). The data imputation method did not affect the results compared to no imputation in the EBM model (Supplementary Materials S11). The percentage of missingness for each variable is reported in Supplementary Figure S12.

## Discussion

### Principal findings

Generalised additive models (GAMs) may on average reduce referrals by about 19% relative to FIT ≥10 µg/g when required to detect the same percentage of cancers as FIT (approximately 90%). This comes at the cost of missing a small proportion of FIT-positive cancers, substituted by extra FIT-negative cancers, since no model reduced referrals while capturing all FIT-positive cancers. However, at higher thresholds where at least 30% of CRCs were captured, GAMs achieved PPVs close to 35%, suggesting that GAM risk scores may be useful for prioritising referred patients to detect about a third of CRCs faster.

### Strengths and limitations

This study used a large sample of 62,219 patients, employed machine learning models that can detect complex cancer signals, and included up to 1,025 clinical predictor variables. Models were optimised for high areas under the ROC and precision-recall curves to ensure that low cancer prevalence did not degrade performance. Performance estimates were obtained through rigorous cross-validation to avoid optimistic estimates: data imputation, normalisation, and variable selection were performed on non-held-out folds. Models were benchmarked against the clinical baseline of FIT ≥10 µg/g, using clinically relevant metrics of referral reduction and referral prioritisation.

The population included adults with a GP FIT and core bloods (HGB, PLT, MCV, WBC), so results may not generalise to patients without bloods, although those with bloods were a majority (80%). In predictor set 3, up to 80% of patients had missing bloods, so learned patterns may not transfer to settings where bloods are ordered for model application. Calibration was assessed minimally, as the focus was on referral reduction and referral prioritisation. The six-month follow-up maximized sample size but likely missed some FIT-negative cancers. However, capturing all FIT-positive cancers is more important as referral reduction and prioritisation are computed relative to FIT-positive patients, and most FIT-positive cancers are expected to be detected within six months given the NHS faster diagnosis targets. Model risk score thresholds for referral reduction were estimated from the test set to place models on an equal cancer detection footing for a clear comparison; real-world use requires estimating the thresholds from training data, which may require an extra data split for unbiased estimation. Finally, the most informative NODE-GAM variables reflect patterns the model used for classification, some of which may not be statistically robust.

### Promising model architectures

Generalised additive models (EBM, NODE-GAM) performed the best, achieving the highest potential to reduce or prioritise referrals. These models balance flexibility (non-linearity) with interpretability (the contribution of each variable can be quantified-visualised).

### Findings in context

CRC risk prediction models have been developed in symptomatic patients – see Hampton et al^2^ and our previous Supplement S17 in Tamm et al^11^. Only three studies^4–6^ have combined FIT with routine data in the full target population of symptomatic primary care patients with FIT results, rather than asymptomatic or referred patients. Withrow et al^4^ found that adding age, sex, and bloods to FIT in logistic models in 16,604 OUH patients did not increase PPV (and thus did not reduce colonoscopies) at the sensitivity of FIT ≥10 µg/g; however, their cohort comprised of patients with lower-risk symptoms, while we analyse a contemporary pan-risk population. Crooks et al^5^ reported a 20% referral reduction with a Cox model in 34,435 Nottingham patients, close to the 16.7% average reduction observed here; but like ML models, COLOFIT achieved no referral reduction if required to capture cancers already detected by FIT ≥10 µg/g. Digby et al^6^ added sex, age, iron deficiency anaemia, and systemic inflammation index to FIT, yet at colonoscopy numbers matching FIT ≥10 µg/g, only three extra CRCs were captured – not substantially outperforming FIT.

The remaining models are less relevant, having not been developed in the full symptomatic FIT primary care population. FAST^32^ and COLONPREDICT^33^ were developed primarily in referred secondary care patients; COLONOFIT^34^ in referred patients with <12% referred due to FIT. Lucocq et al^35^ built ML models in referred primary care patients and reported 30-54% colonoscopy reduction, but did not report cancer miss rate relative to FIT or check whether the reduction remained when compared against a FIT-only model. Models have also been developed in asymptomatic patients, such as COLONFLAG^36^ (age, sex, full blood count and its trends). Given FIT’s strong performance (c-statistic 0.92), models that omit FIT and rely on non-cancer-specific predictors are unlikely to outperform FIT-based models.

### The future of colorectal cancer risk prediction in symptomatic patients

Models may help in two scenarios: (1) reducing referrals, or (2) fast-tracking higher-risk referred patients. Reducing referrals is harder, as a model must capture most cancers (roughly 90%) to match current practice of FIT ≥ 10 µg/g. In this comprehensive comparison, no model reduced referrals if required to capture all cancers detected by FIT ≥10 µg/g. Referral prioritisation appears more promising: GAMs achieved a number needed to scope close to three for at least 30% of colorectal cancers.

Two unexplored model types may improve referral reduction: mixture-of-experts (MoE) models^37^, in which a router model selects the prediction model for each patient, and sequence models that ingest medical events over time (e.g. Wang et al^38^). MoE could help if different subpopulations have different predictor-outcome relationships (e.g. younger vs older). Sequence models can help contextualise inputs (e.g. treating an abnormal blood as relevant only if unexplained by surrounding medical conditions).

The biggest barrier to further progress is probably access to a sufficiently large dataset. In the UK, such data is stored across separate NHS trusts. Given the low prevalence of CRC in the target population (approximately 1%), trust-specific datasets will likely remain small. The new Thames Valley and Surrey Secure Data Environment^39^ is one project that could address this by providing access to a multi-trust dataset.

## Data availability

The data is based on anonymised electronic patient records and cannot be publicly shared.

## Code availability

The code is available at https://github.com/tammandres/fitml

## Funding

The research was supported by the National Institute for Health and Care Research (NIHR) Oxford Biomedical Research Centre. The views expressed are those of the author(s) and not necessarily those of the NHS, the NIHR or the Department of Health. AT was supported by the EPSRC Centre for Doctoral Training in Health Data Science (EP/S02428X/1) during part of the work. BDN was supported by a National Institute of Health and Care Research Academic Clinical Lectureship and a CRUK Research Careers Committee Postdoctoral Fellowship (RCCPDF\100005), and the Oxford Cancer Research UK Centre. EJA was funded by the Oxford Cancer Research UK Centre Bobby Moore Fund (C23434/A23706).

## Competing interests

BDN has received institutional research funding from NIHR, Cancer Research UK and GRAIL; consulting fees from the Multicancer Early Detection (MCED) consortium; and speaker fees from the Royal College of GPs. JEE declares consulting fees from Exact Sciences (now Abbott).

## Acknowledgments

We acknowledge Prof Diana Withrow for a helpful discussion on methods.

## AI use statement

Claude Sonnet 4.6 and Opus 4.8 were used to reduce word count. Claude Code was used to screen code for bugs and reuse existing code for figures. All AI suggestions were manually verified.

## Notes

### Author Declarations

The study was conducted as a service evaluation (Oxford University Hospitals audit number 9076).

## References

1. Booth, R. et al. Role of the faecal immunochemical test in patients with risk-stratified suspected colorectal cancer symptoms: A systematic review and meta-analysis to inform the ACPGBI/BSG guidelines. Lancet Reg. Health – Eur. 23, (2022).

2. Hampton, J. S., et al. The performance of FIT-based and other risk prediction models for colorectal neoplasia in symptomatic patients: a systematic review. eClinicalMedicine 64, (2023).

3. Burnett, B. et al. Machine Learning in Colorectal Cancer Risk Prediction from Routinely Collected Data: A Review. Diagnostics 13, 301 (2023).

4. Withrow, D. R. et al. Combining faecal immunochemical testing with blood test results for colorectal cancer risk stratification: a consecutive cohort of 16,604 patients presenting to primary care. BMC Med. 20, 116 (2022).

5. Crooks, C. J. et al. COLOFIT: Development and Internal-External Validation of Models Using Age, Sex, Faecal Immunochemical and Blood Tests to Optimise Diagnosis of Colorectal Cancer in Symptomatic Patients. Aliment. Pharmacol. Ther. 61, 852–864 (2025).

6. Digby, J. et al. Do risk scores improve use of faecal immunochemical testing for haemoglobin in symptomatic patients in primary care? Colorectal Dis. 26, 675–683 (2024).

7. National Institute for Health and Care Excellence (NICE). Quantitative faecal immunochemical testing to guide colorectal cancer pathway referral in primary care. Diagnostics guidance [DG56]. https://www.nice.org.uk/guidance/dg56 (2023).

8. Beaton, D. et al. UK endoscopy workload and workforce patterns: is there potential to increase capacity? A BSG analysis of the National Endoscopy Database. Frontline Gastroenterol. 14, 103– 110 (2023).

9. Collins, G. S. et al. Protocol for development of a reporting guideline (TRIPOD-AI) and risk of bias tool (PROBAST-AI) for diagnostic and prognostic prediction model studies based on artificial intelligence. BMJ Open 11, e048008 (2021).

10. Tamm, A., Jones, H., Doshi, N., &, et al. Supporting cancer research on real-world data: Extracting colorectal cancer status and explicitly written TNM stages from free-text imaging and histopathology reports (manuscript accepted by BMJ Health & Care Informatics). 10.1136/bmjhci-2025-101521 (2025) doi:10.1136/bmjhci-2025-101521.

11. Tamm, A. et al. External validation of the COLOFIT colorectal cancer risk prediction model in the Oxford-FIT dataset: the importance of population characteristics and clinically relevant evaluation metrics. BMC Med. 23, 503 (2025).

12. Tamm, A. et al. Establishing a colorectal cancer research database from routinely collected health data: the process and potential from a pilot study. BMJ Health Care Inform. 29, e100535 (2022).

13. Friedman, J. H., Hastie, T. & Tibshirani, R. Regularization Paths for Generalized Linear Models via Coordinate Descent. J. Stat. Softw. 33, 1–22 (2010).

14. Breiman, L. Random Forests. Mach. Learn. 45, 5–32 (2001).

15. Chen, T. & Guestrin, C. XGBoost: A Scalable Tree Boosting System. in Proceedings of the 22nd ACM SIGKDD International Conference on Knowledge Discovery and Data Mining 785–794 (Association for Computing Machinery, New York, NY, USA, 2016). doi:10.1145/2939672.2939785.

16. Agarwal, R. et al. Neural Additive Models: Interpretable Machine Learning with Neural Nets. in Advances in Neural Information Processing Systems (eds. Ranzato, M., Beygelzimer, A., Dauphin, Y., Liang, P. S. & Vaughan, J. W.) vol. 34 4699–4711 (Curran Associates, Inc., 2021).

17. Xu, S., Bu, Z., Chaudhari, P. & Barnett, I. J. Sparse Neural Additive Model: Interpretable Deep Learning with Feature Selection via Group Sparsity. in Machine Learning and Knowledge Discovery in Databases: Research Track (eds. Koutra, D., Plant, C., Gomez Rodriguez, M., Baralis, E. & Bonchi, F.) 343–359 (Springer Nature Switzerland, Cham, 2023). doi:10.1007/978-3-031-43418-1_21.

18. Lou, Y., Caruana, R., Gehrke, J. & Hooker, G. Accurate intelligible models with pairwise interactions. in Proceedings of the 19th ACM SIGKDD international conference on Knowledge discovery and data mining 623–631 (Association for Computing Machinery, New York, NY, USA, 2013). doi:10.1145/2487575.2487579.

19. Chang, C.-H., Caruana, R. & Goldenberg, A. NODE-GAM: Neural Generalized Additive Model for Interpretable Deep Learning. in (2021).

20. Grinsztajn, L., Oyallon, E. & Varoquaux, G. Why do tree-based models still outperform deep learning on typical tabular data? Adv. Neural Inf. Process. Syst. 35, 507–520 (2022).

21. Akiba, T., Sano, S., Yanase, T., Ohta, T. & Koyama, M. Optuna: A Next-generation Hyperparameter Optimization Framework. in Proceedings of the 25th ACM SIGKDD International Conference on Knowledge Discovery & Data Mining 2623–2631 (Association for Computing Machinery, New York, NY, USA, 2019). doi:10.1145/3292500.3330701.

22. Davis, J. & Goadrich, M. The relationship between Precision-Recall and ROC curves. in Proceedings of the 23rd international conference on Machine learning 233–240 (Association for Computing Machinery, New York, NY, USA, 2006). doi:10.1145/1143844.1143874.

23. Paszke, A. et al. PyTorch: An Imperative Style, High-Performance Deep Learning Library. In Advances in Neural Information Processing Systems vol. 32 (Curran Associates, Inc., 2019).

24. Qi, Q., Luo, Y., Xu, Z., Ji, S. & Yang, T. Stochastic Optimization of Areas Under Precision-Recall Curves with Provable Convergence. in Advances in Neural Information Processing Systems vol. 34 1752–1765 (Curran Associates, Inc., 2021).

25. Yuan, Z., Yan, Y., Sonka, M. & Yang, T. Large-Scale Robust Deep AUC Maximization: A New Surrogate Loss and Empirical Studies on Medical Image Classification. in 3040–3049 (2021).

26. Pedregosa, F. et al. Scikit-learn: Machine Learning in Python. J. Mach. Learn. Res. 12, 2825–2830 (2011).

27. Simkus, V., Rhodes, B. & Gutmann, M. U. Variational Gibbs Inference for Statistical Model Estimation from Incomplete Data. J. Mach. Learn. Res. 24, 1–72 (2023).

28. The pandas development team. pandas-dev/pandas: Pandas. Zenodo 10.5281/zenodo.10697587 (2024).

29. Nori, H., Jenkins, S., Koch, P. & Caruana, R. InterpretML: A Unified Framework for Machine Learning Interpretability. ArXiv190909223 Cs Stat http://arxiv.org/abs/1909.09223 (2019).

30. Chang, C.-H. (Kingsley). zzzace2000/nodegam. (2024).

31. Yuan, Z. et al. LibAUC: A Deep Learning Library for X-Risk Optimization. in Proceedings of the 29th ACM SIGKDD Conference on Knowledge Discovery and Data Mining 5487–5499 (2023). doi:10.1145/3580305.3599861.

32. Cubiella, J. et al. The fecal hemoglobin concentration, age and sex test score: Development and external validation of a simple prediction tool for colorectal cancer detection in symptomatic patients. Int. J. Cancer 140, 2201–2211 (2017).

33. Cubiella, J. et al. Development and external validation of a faecal immunochemical test-based prediction model for colorectal cancer detection in symptomatic patients. BMC Med. 14, 128 (2016).

34. Fernández-Bañares, F. et al. Prediction of advanced colonic neoplasm in symptomatic patients: a scoring system to prioritize colonoscopy (COLONOFIT study). BMC Cancer 19, 734 (2019).

35. Lucocq, J., Barron, E., Holmes, H., Donnelly, P. D. & Cruickshank, N. Optimising the use of colonoscopy to improve risk stratification for colorectal cancer in symptomatic patients: A decision-curve analysis. Scott. Med. J. 69, 61–71 (2024).

36. Kinar, Y. et al. Development and validation of a predictive model for detection of colorectal cancer in primary care by analysis of complete blood counts: a binational retrospective study. J. Am. Med. Inform. Assoc. 23, 879–890 (2016).

37. Cai, W. et al. A Survey on Mixture of Experts in Large Language Models. IEEE Trans. Knowl. Data Eng. 37, 3896–3915 (2025).

38. Wang, L. et al. Transformer-based deep learning model for the diagnosis of suspected lung cancer in primary care based on electronic health record data. eBioMedicine 110, (2024).

39. Thames Valley and Surrey Secure Data Environment (TVS SDE) – Thames Valley and Surrey. https://thamesvalleyandsurreyhealthandcaredata.nhs.uk/using-patient-data/thames-valley-and-surrey-secure-data-environment/.

