## Supplementary Materials for "Machine learning combining FIT with up to 1,025 clinical variables: limited referral reduction but potential for faster diagnosis"

### S1. Hyperparameter tuning

#### Hyperparameter tuning ranges

Hyperparameter tuning ranges for the models are reported in Table S1-A, and for the alternative loss functions in Table S1-B. These can also be seen in the codebase in /fitml/basemodels.py and /fitml/params.py.

For models implemented in pytorch, additional training parameters were tuned. Learning rate was tuned in [1e-3, 2e-2] during primary training and in [1e-4, 1e-2] during finetuning, using log-uniform sampling. When fine-tuning with ap, aucm and caucm losses, proportion of positive samples in each batch was tuned in [0.01, 0.1, 0.3, 0.5]. When training with bce-hardpos loss, proportion of positive samples was tuned in [0.03, 0.05, 0.1].

For all non-tree based models, data transform was tuned in ['norm', 'logstandard'], and sample weight ratio was tuned in [None, 2, 4]. The sample weight ratio indicates how much the weight given to a newer primary subset of data is higher compared to an older subset of data (see "Incorporating non-buffered data" in Methods for motivation); if None, then no sample weights are used. Sample weight ratio was not applied when fine-tuning pretrained models with alternative loss functions (ap, aucm, caucm), because it was not straightforward to apply sample weights in the existing python package that provided the alternative losses.

**Table S1-A**. Hyperparameter tuning ranges for machine learning models

| Model | Parameter | Sampling | Low | High | Step | Choices |
|---|---|---|---|---|---|---|
| **Pytorch-models** | | | | | | |
| MLP | dropout | uniform | 0 | 0.5 | | |
| MLP | l1_lambda | log-uniform | 1.00E-05 | 1000 | | |
| MLP | l2_lambda | log-uniform | 1.00E-05 | 1000 | | |
| MLP | skip | categorical | | | | False; True |
| NAM | arch | categorical | | | | 64-64-32 |
| NAM | feature_dropout | uniform | 0 | 0.2 | | |
| NAM | hidden_dropout | uniform | 0 | 0.5 | | |
| NAM | l2_lambda | log-uniform | 1.00E-05 | 1000 | | |
| NAM | linear_l1 | log-uniform | 1.00E-05 | 100 | | |
| NAM | linear_l2 | log-uniform | 1.00E-05 | 100 | | |
| NAM | out_l2 | log-uniform | 1.00E-05 | 100 | | |
| NODE-GAM | colsample_bytree | categorical | | | | 1e-05; 0.1; 0.5; 1 |
| NODE-GAM | depth | categorical | | | | 2; 4 |
| NODE-GAM | last_dropout | discrete-uniform | 0 | 0.8 | 0.1 | |
| NODE-GAM | num_layers | categorical | | | | 2; 3; 4 |
| NODE-GAM | num_trees | categorical | | | | 50; 100; 200; 400; 1000 |
| NODE-GAM | out_l2 | categorical | | | | 0.0; 1e-07; 1e-06; 1e-05 |
| NODE-GAM | output_dropout | discrete-uniform | 0 | 0.5 | 0.1 | |
| PLR* | C | log-uniform | 0.1 | 10000 | | |
| PLR | l1_ratio | uniform | 0 | 1 | | |
| SNAM | group_l1 | log-uniform | 1.00E-05 | 100 | | |
| SNAM | linear_l1 | log-uniform | 1.00E-05 | 100 | | |
| SNAM | linear_l2 | log-uniform | 1.00E-05 | 100 | | |
| **Base ML models** | | | | | | |
| EBM | early_stopping_rounds | categorical | | | | 10; 100 |
| EBM | interactions | int-uniform | 0 | 20 | 5 | |
| EBM | learning_rate | log-uniform | 0.001 | 0.1 | | |
| EBM | max_bins | categorical | | | | 32; 64; 128; 1024 |
| EBM | max_interaction_bins | categorical | | | | 4; 32; 64 |
| EBM | max_leaves | categorical | | | | 2; 3 |
| EBM | reg_alpha | log-uniform | 0.001 | 100 | | |
| EBM | reg_lambda | log-uniform | 0.001 | 100 | | |
| EBM | validation_size | categorical | | | | 0.15; 0.3 |
| RF | max_depth | int-uniform | 2 | 12 | 1 | |
| RF | max_features | uniform | 0.4 | 1 | | |
| RF | min_samples_leaf | int-uniform | 1 | 100 | 1 | |
| RF | min_samples_split | int-uniform | 2 | 100 | 1 | |
| XGB | colsample_bytree | uniform | 0.4 | 1 | | |
| XGB | gamma | log-uniform | 0.001 | 10 | | |
| XGB | learning_rate | log-uniform | 0.001 | 0.3 | | |
| XGB | max_depth | int-uniform | 3 | 8 | 1 | |
| XGB | min_child_weight | int-uniform | 1 | 100 | 1 | |
| XGB | reg_alpha | log-uniform | 1.00E-05 | 1000 | | |
| XGB | reg_lambda | log-uniform | 1.00E-05 | 1000 | | |
| XGB | subsample | uniform | 0.7 | 1 | | |

*Notes*. Models: PLR - penalised logistic regression, EBM - explainable boosting classifier, NAM - neural additive model, SNAM - sparse neural additive model, NODE-GAM - neural oblivious decision tree ensemble GAM, RF - random forests, XGB - XGBoost, MLP - multilayer perceptron. *The PLR model was implemented in pytorch, so the tuning range for C and l1_ratio will not necessarily match that of the scikit-learn's implementation.

**Table S1-B**. Hyperparameter tuning ranges for alternative loss functions

| Loss | Parameter | Sampling | Low | High | Step | Choices |
|---|---|---|---|---|---|---|
| aucm | auc_margin | categorical | | | | 0.1; 0.3; 0.5; 0.7; 1; 2; 5; 10 |
| aucm | auc_epoch_decay | categorical | | | | 0; 1e-06; 1e-05; 0.0001; 0.001 |
| ap | ap_margin | categorical | | | | 0.1; 0.3; 0.5; 0.7; 1; 2; 5; 10 |
| ap | ap_gamma | categorical | | | | 0.1; 0.3; 0.5; 0.7; 0.9; 0.99 |
| bce-hardpos | hardpos_alpha | uniform | 0.5 | 0.95 | | |
| bce-hardpos | hardpos_margin | uniform | 0.1 | 0.4 | | |

*Notes.* aucm - auc margin loss, caucm - compositional auc margin loss, ap - average precision loss, bce-hardpos - binary-cross entropy loss with hard-positive penalty

#### Additional details of hyperparameter tuning

Some models were implemented in pytorch: PLR, NAM, SNAM, NODE-GAM, and MLP (Table 2). In each cross-validation fold, 50 hyperparameter configurations (trials) were evaluated for each model. This gives a total of 500 hyperparameter evaluations per model, because there were ten folds in total (five-fold cross-validation repeated twice). To evaluate each parameter combination, a model was trained for at most 100 epochs (the data was split into minibatches - one cycle through all minibatches constitutes one epoch). To speed up the process, early stopping and pruning were used (see below). Further, for models trained on GPU (NAM, SNAM, NODE-GAM), two hyperparameter trials were run in parallel for an additional speed-up.

**Generation of hyperparameter configurations**. Hyperparameter configurations were sampled using tree-structured parzen (TPE) estimators, a method that suggests the next hyperparameter combination based on the performance of previously evaluated configurations. The first ten hyperparameter configurations were sampled randomly and the remaining 40 using the TPE algorithm. At each step, the algorithm ranks currently completed trials based on validation set scores, divides them into higher- and lower-ranked groups, and uses kernel density estimation to estimate the distribution of each parameter in the two groups. TPE then suggests the next parameter combination, such that for each parameter value, it maximizes the probability density under the distribution of higher-ranked trials and minimizes the density under the distribution of lower-ranked trials. TPE was used to intelligently suggest the next hyperparameter combination, so that a higher performing combination could be found given the limited number of tuning trials.

**Learning rate decay.** The models were trained for at most 100 epochs using an exponential learning rate decay with gamma = 0.97625, which ensured that the learning rate decayed to approximately 10% of the starting learning rate by epoch 100 (0.97625^100 = 0.09).

**Early stopping**. During tuning, the mean validation set score across the five most recent epochs was compared to the mean validation set score of the earlier five epochs. If these scores did not improve for three consecutive epochs, tuning was stopped. Early stopping was activated after 1000 parameter update steps (approximately 19 epochs) for the NODE-GAM model, and after 500 update steps for other models. A higher warm-up period was used for NODE-GAM, because the model had to be trained for a minimum of 2000 steps for the temperature annealing mechanism to converge (which ensures that the model becomes an interpretable generalised additive model). For NODE-GAM, early stopping from step 1000 onwards was a compromise, allowing less promising configurations to be terminated earlier, while ensuring that a substantial number of optimisation steps have occurred to meaningfully evaluate the current parameter combination.

**Pruning**. During tuning, a percentile pruner was applied after the models had completed training for 20 epochs. At each subsequent epoch, the pruner terminated trials that were in the bottom 25% of validation set scores among all trials. The 25-th percentile rather than the median was used to make the pruner less aggressive. The 20-epoch warmup period was chosen to ensure that roughly 1000 optimisation steps will always be completed for the NODE-GAM model that requires a minimum of 2000 steps to be fully trained. Therefore, a tuning trial could be stopped early due to the validation set score not improving across training steps (the early stopping mechanism described above), or the trial performing worse than existing trials at the current step (the pruning mechanism).

#### S2. Optional sample weights for secondary (non-buffered) FIT data

The dataset consisted of a primary subset (patients tested with buffered FIT test kits) and a secondary subset (patients not tested with buffered kits) - see "Incorporating non-buffered data". To ensure that the models reflect current practice, only primary subset was used for model evaluation and was divided into cross-validation folds. To make use of information contained in the secondary subset, the secondary subset was added to the training data in each fold, but the samples were optionally down-weighted (the weight was selected during tuning, including the possibility of no weight).

The sample weights for the primary subset (w1) and the secondary subset (w0) were defined as

$$w_0 = \frac{n}{n_0(1+r)}, \quad w_1 = \frac{r \cdot n}{n_1(1+r)}$$

where $w_0$ - weight applied to each sample in the secondary subset; $w_1$ - weight applied to each sample in the primary subset; $n_0$ – number of samples in the secondary subset; $n_1$ - number of samples in the primary subset; $r$ – "sample weight ratio", the ratio of the total weight applied to the primary subset and the secondary subset.

The weights were computed using these formulas, because then (1) the sample weight ratio is the ratio of total weights applied to each data subset, and (2) the average weight is equal to one.

Condition (1) can be seen from:

$$\frac{n_1 w_1}{n_0 w_0} = r$$

due to

$$n_0 w_0 = \frac{n}{1+r}, \qquad n_1 w_1 = \frac{rn}{1+r}$$

And condition (2) can be seen from:

$$\frac{(n_0 w_0 + n_1 w_1)}{n} = \frac{\left(\frac{n}{1+r} + \frac{rn}{1+r}\right)}{n} = 1$$

The initial weighting scheme was suggested by Claude AI, then verified-checked by the first author.

#### S3. Descriptive statistics for selected bloods in the primary subset

**Table S3:** Summary of selected blood tests for the patient cohort in the primary subset of patients with buffered test kits

| | No colorectal cancer | Colorectal cancer | Statistical test |
|---|---|---|---|
| Number of patients | 30547 | 315 | |
| Haemoglobin (HGB) | | | |
| Median (25th, 75th) | 133.0 (121.0, 144.0) | 128.0 (109.5, 140.5) | |
| Min, max | 39.0, 221.0 | 68.0, 185.0 | |
| Low haemoglobin | 8574 (28.1%) | 150 (47.6%) | X=58.8, p=0.000*** |
| Normal haemoglobin | 21950 (71.9%) | 165 (52.4%) | X=58.2, p=0.000*** |
| Not known | 23 (0.1%) | - | |
| Platelets (PLT) | | | |
| Median (25th, 75th) | 270.0 (225.0, 322.0) | 308.0 (253.5, 373.5) | |
| Min, max | 12.0, 1017.0 | 79.0, 651.0 | |
| High platelets | 2342 (7.7%) | 58 (18.4%) | X=50.2, p=0.000*** |
| Normal platelets | 28185 (92.3%) | 257 (81.6%) | X=49.2, p=0.000*** |
| Not known | 20 (0.1%) | - | |
| White cells (WBC) | | | |
| Median (25th, 75th) | 6.8 (5.6, 8.2) | 7.4 (6.2, 9.0) | |
| Min, max | 0.8, 192.2 | 2.9, 24.6 | |
| High white cells | 1606 (5.3%) | 28 (8.9%) | X=8.2, p=0.004** |
| Normal white cells | 28921 (94.7%) | 287 (91.1%) | X=7.8, p=0.005** |
| Not known | 20 (0.1%) | - | |
| Mean cell haemoglobin (MCH) | | | |
| Median (25th, 75th) | 30.2 (28.7, 31.4) | 29.2 (26.6, 30.9) | |
| Min, max | 12.5, 48.2 | 15.4, 38.7 | |
| Low mean cell haemoglobin | 4197 (13.7%) | 94 (29.8%) | X=67.5, p=0.000*** |
| Normal mean cell haemoglobin | 26328 (86.2%) | 221 (70.2%) | X=66.6, p=0.000*** |
| Not known | 22 (0.1%) | - | |
| Mean cell volume (MCV) | | | |
| Median (25th, 75th) | 91.9 (88.1, 95.5) | 89.9 (84.8, 94.2) | |
| Min, max | 49.5, 141.6 | 58.5, 121.6 | |
| Low mean cell volume | 1549 (5.1%) | 50 (15.9%) | X=74.1, p=0.000*** |
| Normal mean cell volumne | 28978 (94.9%) | 265 (84.1%) | X=72.3, p=0.000*** |
| Not known | 20 (0.1%) | - | |
| Serum ferritin (CFER) | | | |
| Median (25th, 75th) | 69.0 (24.8, 148.7) | 37.0 (13.2, 87.7) | |
| Min, max | 1.0, 32461.3 | 2.7, 567.2 | |
| High serum ferritin | 1059 (3.5%) | 11 (3.5%) | X=0.0, p=0.981 |
| Low serum ferritin | 3435 (11.2%) | 72 (22.9%) | X=41.7, p=0.000*** |
| Normal serum ferritin | 16872 (55.2%) | 185 (58.7%) | X=1.5, p=0.214 |
| Not known | 13675 (44.8%) | 130 (41.3%) | X=1.5, p=0.214 |
| C-reactive protein (CRP) | | | |
| Median (25th, 75th) | 2.1 (0.8, 6.0) | 4.8 (1.3, 13.2) | |
| Min, max | 0.2, 441.4 | 0.2, 267.1 | |
| High C-reactive protein | 3349 (11.0%) | 60 (19.0%) | X=20.7, p=0.000*** |
| Normal C-reactive protein | 16614 (54.4%) | 131 (41.6%) | X=20.6, p=0.000*** |
| Not known | 10643 (34.8%) | 125 (39.7%) | X=3.2, p=0.073 |

Notes. Normal, high, and low values for these bloods were defined as in Withrow et al [10]. Low HGB: < 130 g/L for males, < 120 g/L for females. High PLT: >400 * 109/L. High WBC: > 11 * 109/L. Low MCH: < 27.4 pg/cell. Low MCV: < 80 fl. Low CFER: < 20 µg/L. High CFER: ≥ 350 µg/L. High CRP: > 10 mg/L.

#### S4. Descriptive statistics for the entire patient cohort

Table S4 reports key characteristics for individuals with and without colorectal cancer (CRC) in the entire dataset, where both buffered (primary subset) and non-buffered FIT test kits (secondary subset) were used. The machine learning models were trained on the entire dataset, but optionally giving less weight to patients who had non-buffered test kits.

**Table S4.** Descriptive statistics for the OUH-FIT dataset, covering both non-buffered and buffered FIT test kits

| | No colorectal cancer | Colorectal cancer | Statistical test |
|---|---|---|---|
| Number of patients | 61473 | 746 | X^2(1)=nan, p=nan |
| Age | | | |
| 18-39.9 | 7962 (13.0%) | 20 (2.7%) | X^2(1)=69.5, p<0.001*** |
| 40-49.9 | 8274 (13.5%) | 50 (6.7%) | X^2(1)=29.0, p<0.001*** |
| 50-59.9 | 11558 (18.8%) | 96 (12.9%) | X^2(1)=17.0, p<0.001*** |
| 60-69.9 | 10563 (17.2%) | 124 (16.6%) | X^2(1)=0.2, p=0.686 |
| 70-79.9 | 12631 (20.5%) | 226 (30.3%) | X^2(1)=42.7, p<0.001*** |
| ≥80 | 10485 (17.1%) | 230 (30.8%) | X^2(1)=98.1, p<0.001*** |
| Median (25th, 75th percentile) | 62.0 (49.0, 76.0) | 74.0 (61.0, 81.0) | |
| Min, max | 18.0, 103.0 | 27.0, 102.0 | |
| Gender | | | |
| Female or not specified (<10) | 36474 (59.3%) | 322 (43.2%) | X^2(1)=79.7, p<0.001*** |
| Male | 24999 (40.7%) | 424 (56.8%) | X^2(1)=79.7, p<0.001*** |
| Ethnicity | | | |
| Asian | 1698 (2.8%) | Not Available | |
| Black | 578 (0.9%) | Not Available | |
| Mixed | 441 (0.7%) | Not Available | |
| Other Ethnic Groups | 695 (1.1%) | Not Available | |
| White | 44000 (71.6%) | 545 (73.1%) | X^2(1)=0.8, p=0.373 |
| Not stated | 12244 (19.9%) | 178 (23.9%) | X^2(1)=7.2, p=0.007** |
| Not known | 1817 (3.0%) | Not Available | |
| IMDD | | | |
| Median (25th, 75th percentile) | 8.0 (7.0, 10.0) | 8.0 (7.0, 10.0) | |
| Min, max | 1.0, 10.0 | 1.0, 10.0 | |
| Not known | 9891 (16.1%) | 49 (6.6%) | X^2(1)=49.8, p<0.001*** |
| FIT (µg Hb/g) | | | |
| 0-1.9 | 46440 (75.5%) | 52 (7.0%) | X^2(1)=1835.1, p<0.001*** |
| 2-9.9 | 7486 (12.2%) | 43 (5.8%) | X^2(1)=28.5, p<0.001*** |
| 10-99.9 | 5130 (8.3%) | 230 (30.8%) | X^2(1)=473.4, p<0.001*** |
| ≥100 | 2417 (3.9%) | 421 (56.4%) | X^2(1)=4667.1, p<0.001*** |
| Median (25th, 75th percentile) | 0.0 (0.0, 1.7) | 165.5 (32.4, 400.0) | |
| Min, max | 0.0, 400.0 | 0.0, 400.0 | |
| Symptoms - GP reported | | | |
| Abdominal mass | 78 (0.1%) | Not Available | |
| Abdominal pain | 8077 (13.1%) | 75 (10.1%) | X^2(1)=6.2, p=0.013* |
| Anaemia | 8910 (14.5%) | 155 (20.8%) | X^2(1)=23.4, p<0.001*** |
| Bloating | 2109 (3.4%) | 10 (1.3%) | X^2(1)=9.8, p=0.002** |
| Blood in stool | 7038 (11.4%) | 131 (17.6%) | X^2(1)=27.0, p<0.001*** |
| Change in bowel habit | 13713 (22.3%) | 171 (22.9%) | X^2(1)=0.2, p=0.688 |
| Constipation | 2133 (3.5%) | 13 (1.7%) | X^2(1)=6.6, p=0.010* |
| Diarrhoea | 6561 (10.7%) | 60 (8.0%) | X^2(1)=5.4, p=0.021* |
| Family history of colorectal cancer | 511 (0.8%) | - | |
| Fatigue | 1156 (1.9%) | Not Available | |
| Inflammation | 787 (1.3%) | Not Available | |
| Iron deficiency anaemia | 4179 (6.8%) | 77 (10.3%) | X^2(1)=14.4, p<0.001*** |
| Low iron | 2367 (3.9%) | 19 (2.5%) | X^2(1)=3.4, p=0.065 |
| Melaena | 604 (1.0%) | Not Available | |
| Rectal bleeding | 4382 (7.1%) | 81 (10.9%) | X^2(1)=15.4, p<0.001*** |
| Rectal mass | 33 (0.1%) | Not Available | |
| Rectal pain | 337 (0.5%) | Not Available | |
| Thrombocytosis | 765 (1.2%) | Not Available | |
| Weight loss | 4715 (7.7%) | 59 (7.9%) | X^2(1)=0.1, p=0.808 |
| Not known | 8849 (14.4%) | 99 (13.3%) | X^2(1)=0.8, p=0.384 |
| Haemoglobin (HGB) | | | |
| median (25th, 75th percentile) | 133.0 (120.0, 144.0) | 125.0 (108.2, 140.0) | X^2(1)=50728.3, p<0.001*** |
| min and max | 39.0, 226.0 | 53.0, 185.0 | |
| low haemoglobin | 18246 (29.7%) | 386 (51.7%) | X^2(1)=171.0, p<0.001*** |
| normal haemoglobin | 43192 (70.3%) | 360 (48.3%) | X^2(1)=169.9, p<0.001*** |
| Not known | 35 (0.1%) | - | |
| Platelets (PLT) | | | |
| median (25th, 75th percentile) | 268.0 (224.0, 320.0) | 305.0 (246.2, 371.8) | |
| min and max | 8.0, 1241.0 | 79.0, 920.0 | |
| high platelets | 4688 (7.6%) | 140 (18.8%) | X^2(1)=127.8, p<0.001*** |
| normal platelets | 56754 (92.3%) | 606 (81.2%) | X^2(1)=125.9, p<0.001*** |
| Not known | 31 (0.1%) | - | |
| White cells (WBC) | | | |
| median (25th, 75th percentile) | 6.8 (5.6, 8.2) | 7.4 (6.2, 9.1) | X |
| min and max | 0.8, 286.4 | 2.9, 24.6 | |

| | | | |
|---|---|---|---|
| high white cells | 3204 (5.2%) | 71 (9.5%) | X^2(1)=27.4, p<0.001*** |
| normal white cells | 58238 (94.7%) | 675 (90.5%) | X^2(1)=26.5, p<0.001*** |
| Not known | 31 (0.1%) | - | |
| Mean cell haemoglobin (MCH) | | | |
| median (25th, 75th percentile) | 30.1 (28.6, 31.3) | 28.7 (25.9, 30.6) | |
| min and max | 12.5, 49.6 | 12.5, 38.7 | |
| low mean cell haemoglobin | 8962 (14.6%) | 250 (33.5%) | X^2(1)=209.5, p<0.001*** |
| normal mean cell haemoglobin | 52478 (85.4%) | 496 (66.5%) | X^2(1)=207.7, p<0.001*** |
| Not known | 33 (0.1%) | - | |
| Mean cell volume (MCV) | | | |
| median (25th, 75th percentile) | 91.5 (87.8, 95.0) | 89.0 (83.0, 93.5) | |
| min and max | 49.5, 141.6 | 55.0, 121.6 | |
| low mean cell volume | 3395 (5.5%) | 132 (17.7%) | X^2(1)=204.2, p<0.001*** |
| normal mean cell volumne | 58047 (94.4%) | 614 (82.3%) | X^2(1)=200.9, p<0.001*** |
| Not known | 31 (0.1%) | - | |
| Serum ferritin (CFER) | | | |
| median (25th, 75th percentile) | 69.0 (24.5, 149.3) | 28.8 (12.3, 93.6) | |
| min and max | 1.0, 32461.3 | 1.0, 789.6 | |
| high serum ferritin | 2038 (3.3%) | 23 (3.1%) | X^2(1)=0.1, p=0.725 |
| low serum ferritin | 7022 (11.4%) | 176 (23.6%) | X^2(1)=106.7, p<0.001*** |
| normal serum ferritin | 33239 (54.1%) | 424 (56.8%) | X^2(1)=2.3, p=0.132 |
| Not known | 28234 (45.9%) | 322 (43.2%) | X^2(1)=2.3, p=0.132 |
| C-reactive protein (CRP) | | | |
| median (25th, 75th percentile) | 2.1 (0.9, 6.0) | 5.3 (1.6, 18.1) | |
| min and max | 0.2, 441.4 | 0.2, 267.1 | |
| high C-reactive protein | 6924 (11.3%) | 165 (22.1%) | X^2(1)=86.0, p<0.001*** |
| normal C-reactive protein | 35177 (57.2%) | 314 (42.1%) | X^2(1)=68.9, p<0.001*** |
| Not known | 19480 (31.7%) | 269 (36.1%) | X^2(1)=6.5, p=0.011* |
| CRC-relevant treatments | | | |
| No treatments recorded | 53279 (86.7%) | 109 (14.6%) | X^2(1)=3142.5, p<0.001*** |
| chemotherapy | 1947 (3.2%) | 320 (42.9%) | X^2(1)=3313.5, p<0.001*** |
| local excision | 69 (0.1%) | 29 (3.9%) | X^2(1)=668.0, p<0.001*** |
| radical resection | 332 (0.5%) | 440 (59.0%) | X^2(1)=20543.1,p<0.001*** |
| radiotherapy | 1493 (2.4%) | 120 (16.1%) | X^2(1)=544.4, p<0.001*** |
| T stage | | | |
| 0 | - | Not Available | |
| 1 | - | 77 (10.3%) | |
| 2 | - | Not Available | |
| 3 | - | 226 (30.3%) | |
| 4 | - | 97 (13.0%) | |
| Not known | - | 274 (36.7%) | |

Notes. *CRC-relevant treatments are procedures used for treating colorectal cancer (CRC), but they may also be given for other conditions. **T-stage was extracted from radiology and pathology reports using a pattern-matching algorithm. "Not available" means that the count was < 10, in which case the category with the second lowest count was hidden too so that it could not be derived from the total; this is to comply with the OUH data reporting requirements. Normal, high, and low values for the reported blood test were defined as in Withrow et al [10]. Low HGB: < 130 g/L for males, < 120 g/L for females. High PLT: >400 * 109/L. High WBC: > 11 * 109/L. Low MCH: < 27.4 pg/cell. Low MCV: < 80 fl. Low CFER: < 20 μg/L. High CFER: ≥ 350 μg/L. High CRP: > 10 mg/L.

#### S5. Comparison of patient prioritisation strategies

**Motivation.** The NODE-GAM model achieved approximately 35% PPV at 30% sensitivity (versus 14.6% PPV of FIT), indicating a potential to detect a third of CRCs faster. To explore this further, a simulation was run to compare four strategies of prioritising FIT-positive patients for colorectal investigation:

(1) Patients with higher NODE-GAM risk scores are investigated first;
(2) Patients with higher COLOFIT risk scores are investigated first;
(3) Patients with higher FIT values are investigated first;
(4) FIT-positive patients are investigated in a random order.

For simplicity, it was assumed that all FIT-positive patients will be investigated sequentially – a reasonable approximation if patients in a colonoscopy centre queue for a single colonoscopy slot.

**Method.** In each of the ten held-out cross-validation (CV) folds, patients were ordered by strategies 1-4. For each strategy, the percentage of CRCs detected was recorded against the percentage of patients investigated (those higher in the order first). NODE-GAM and COLOFIT scores were unique per patient, so strategies (1) and (2) provided a single ordering per fold. For strategies (3) and (4), 500 random patient orderings were generated per fold; under strategy (3), these differed only at positions where patients had the same FIT value. We then average the percentage of CRCs detected at each percentage investigated across the 500 random orderings. Finally, to aggregate results across CV folds, the per-fold curves of percent investigated vs percent detected were linearly interpolated onto a common grid of percent investigated at 1% increments (1%, 2%, …, 99%, 100% investigated). An average percentage of cancers detected across the CV folds was reported along with plus-minus one standard deviation to indicate variability across folds.

**Results.** By the time first 5% of patients had been sequentially investigated, NODE-GAM scores led to the identification of 30.5% (SD = 5.25%) of CRCs and FIT values to 13% (SD = 1.5%) of CRCs on average (Figure S5). Of the CRCs detected among the top 5% of patients prioritised by NODE-GAM, 49.3% subsequently had a pathological T-stage of 3 or 4 (locally advanced tumour), and 37.8% had an unknown pathological T-stage. The percentage of patients with a known T-stage of 3 or 4 remained similar across the different percent of patients investigated, and was slightly higher in patients investigated first.

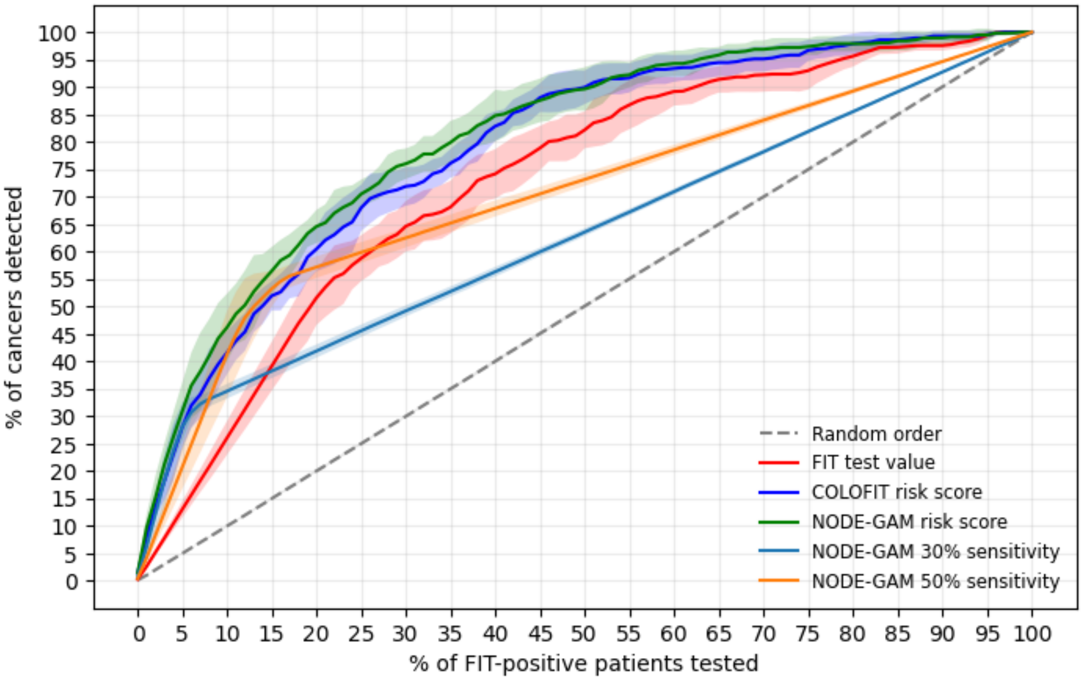

**Figure S5**. **Cumulative percent cancers detected per percent patients investigated under different patient prioritisation strategies.** The simulation assumes that patients with a positive FIT test value (FIT ≥10 µg/g) are sequentially tested in an order defined by one of the strategies. Strategies: NODE-GAM risk score – patients with higher NODE-GAM model risk score are investigated first; COLOFIT risk score – patients with higher COLOFIT model risk score are investigated first; FIT value – patients with higher FIT test values are investigated first; NODE-GAM 30% sensitivity - patients above the NODE-GAM risk score that captures 30% of FIT-positive cancers are investigated first; NODE-GAM 50% sensitivity – patients above the NODE-GAM risk score threshold that captures 50% of FIT-positive cancers are investigated first. For strategies that did not yield a unique ordering of patients, 500 randomly sampled patient orderings were created, and the average percent of cancers detected per each percent of patients tested was computed. The figure show an average percent of cancers detected across the ten cross-validation folds; the shaded area shows plus-minus one standard deviation across the folds. Per-fold data was linearly interpolated to a common grid of “percent patients tested” with 1% step (1%, 2%, …, 99% tested) to aggregate it across folds.

#### S6. ROC, PR and test-reduction curves with uncertainty intervals

Figure S6 shows the average ROC, precision-recall, and test-reduction curve for each model over the cross-validation folds, and the shaded area shows plus-minus one standard deviation across the cross-validation folds. Standard errors were not used, as they are not straightforward to compute, given the non-independence of the estimates in different folds (as the estimates are derived from models that have been trained on overlapping sets of data).

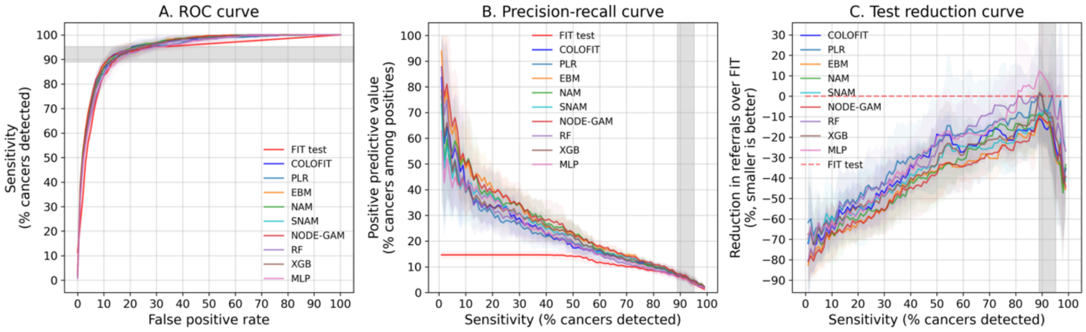

**Figure S6. Receiver-operating characteristic (ROC), precision-recall and test-reduction curves for machine learning models, COLOFIT and the FIT test**. Curves show the mean value of each quantity over ten held-out cross-validation folds; shaded areas show plus-minus one standard deviation. The vertical grey area is shaded between the minimum and maximum sensitivities of FIT ≥10 µg/g in the held-out folds. The test reduction curve shows the percent reduction in the number of positive tests relative to FIT at each level of sensitivity (the model and FIT thresholds yield the same sensitivity). All curves were interpolated to a fixed grid of values at 1-unit increments. Abbreviations: FIT – faecal immunochemical test, COLOFIT – an existing prediction model, PLR – penalised logistic regression, EBM – explainable boosting machine, NAM – neural additive model, SNAM – sparse neural additive model, RF – random forests, XGB – XGBoost, MLP – multilayer perceptron.

#### S7. Feature importance curves by cross-validation fold

Generalised additive models permit examining the contribution of individual variables via feature importance scores and shape graphs. For both EBM and NODE-GAM, the top ten most important variables across the cross-validation folds were FIT, age, sex, change in bowel habit (yes/no), changes in MCV and mean cell haemoglobin (MCH) over a 3-year period before FIT, PLT, albumin, serum ferritin, and the urea blood test order (yes/no). The shape graphs of these variables reveal patterns (Figure S7-A illustrates NODE-GAM shape graphs). First, an increase in FIT from zero to roughly 10 µg/g was associated with a faster increase in CRC risk than between 10 and 400 µg/g. Secondly, age had an inverted U-shaped association with CRC. Thirdly, a decline in MCV and MCH over time was associated with increased CRC risk. Fourthly, increased PLT was associated with CRC within the normal range (150 – 450 10^9/L). Figure S7-A displays the 'average' shape graph across CV folds; per-fold graphs can be found in Figure S7-B. These associations were picked up by ML models and not statistically tested – See Discussion.

In each fold, NODE-GAM was trained on top 100 variables selected by XGBoost. The top 20 most important NODE-GAM variables across folds were defined as those which (1) were selected by XGBoost in at least five folds, and (2) had the highest mean NODE-GAM feature importance score across folds. The feature importance in this case is calculated by first binning the continuous values of a variable into at most 256 bins, then computing the variable's centered contribution to the linear predictor per bin, and finally computing a weighted average of the absolute values of these contributions across bins such that the weights are proportional to the number of observations in each bin. Intuitively, this means that if the shape graph tends to be high above zero in some regions and low below zero in other regions of the input variable, the feature importance is higher, and especially if these regions have a large number of patients. Conversely, if some values of the variable are associated with a very large contribution to the linear predictor (positive or negative), but there are only a few patients with these values, then this contribution is considered less (is down-weighted) when computing feature importance.

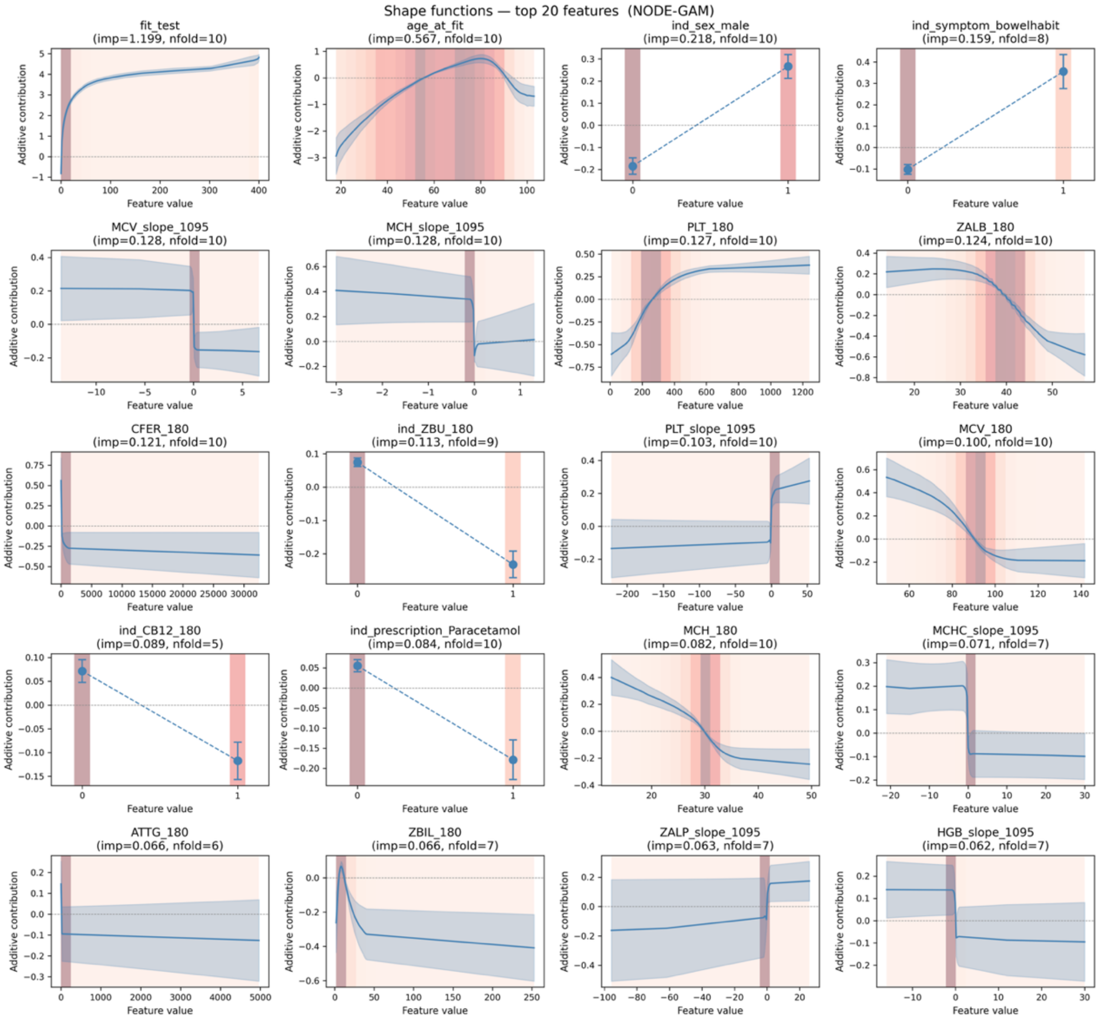

**Figure S7-A.** Top 20 most informative variables for predicting colorectal cancer (CRC) according to the NODE-GAM generalised additive model. Shape graphs show the mean ±1 standard deviation across the ten cross-validation folds. In each fold, NODE-GAM was trained on the top 100 variables selected by XGBoost for that fold. The top 20 variables shown were selected by XGBoost in at least five folds and had the highest mean feature importance score across folds. Shaded vertical bars are histogram bins (darker colour - higher proportion of patients). Variable names: fit_test – faecal immunochemical test (FIT), age_at_fit – age at FIT testing, ind_sex_male – indicator for male sex, ind_symptom_bowelhabit – indicator for change in bowel habit, MCV_slope_1095 – slope of the mean cell volume (MCV) time series over a 1905 day (3-year) period before FIT, MHC_slope_1095 – slope of the mean cell haemoglobin (MCH) time series, PLT_180 – most recent value of platelets (PLT) in a 180-day window before FIT, ZALB_180 – albumin, CFER_180 – serum ferritin, ind_ZBU_180 – indicator for urea test, PLT_slope_1095 – slope of the platelets time series, MCV_180 – mean cell volume, ind_CB12_180 – indicator for vitamin B12 test, ind_prescription_paracetamol – indicator for paracetamol prescription, MCH_180 – mean cell haemoglobin, MCHC_slope_1095 – slope of the mean cell haemoglobin concentration time series, ATTG_180 – anti-tissue transglutaminase, ZBIL_180 – bilirubin, ZALP_slope_1095 – slope of alkaline phosphatase time series, HGB_slope_1095 – slope of haemoglobin time series.

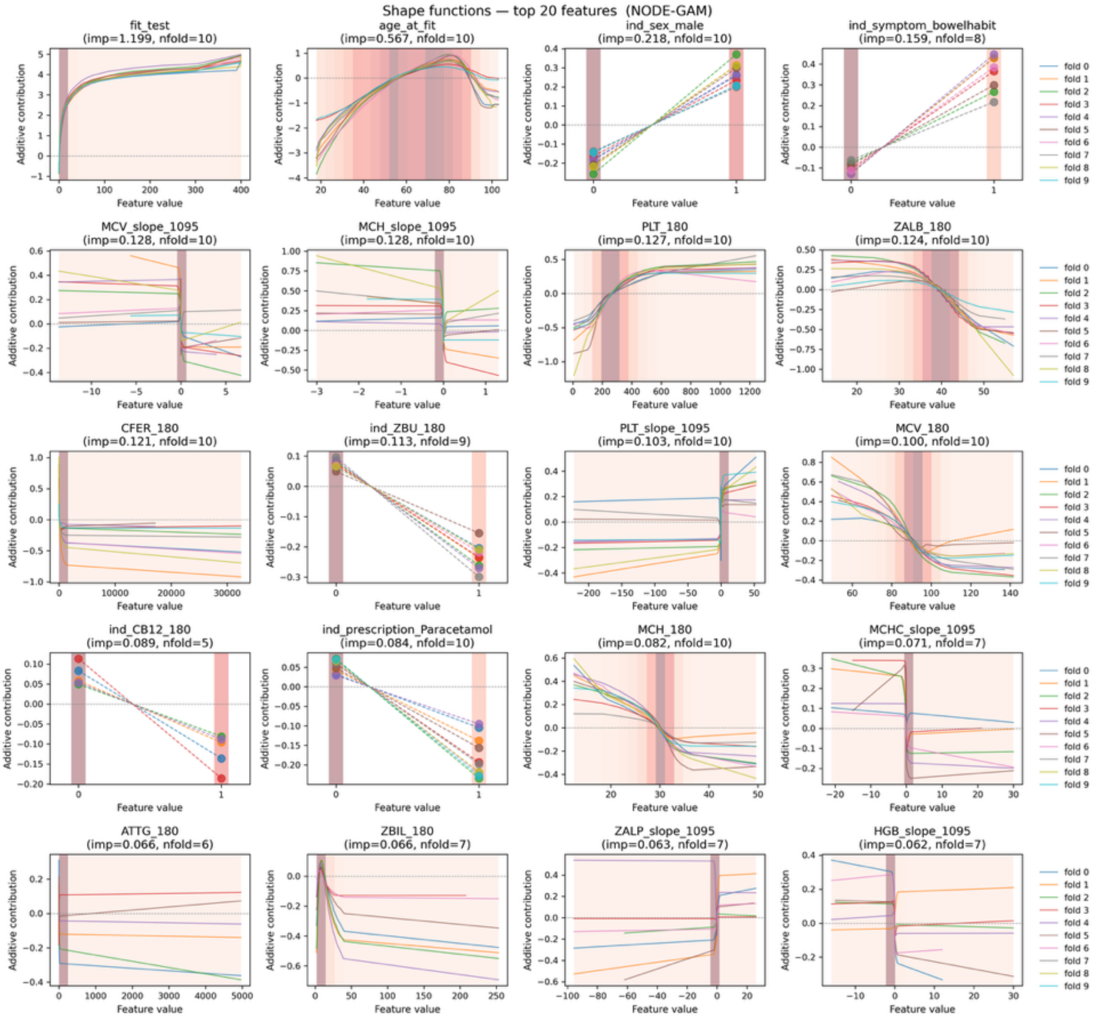

**Figure S7-B**. Top 20 most informative variables for predicting colorectal cancer (CRC) according to the NODE-GAM generalised additive model – per fold shape graphs. In each fold, NODE-GAM was trained on the top 100 variables selected by XGBoost for that fold. The top 20 variables shown were selected by XGBoost in at least five folds and had the highest mean feature importance score across folds. Shaded vertical bars are histogram bins (darker colour - higher proportion of patients). Variable names: fit_test – faecal immunochemical test (FIT), age_at_fit – age at FIT testing, ind_sex_male – indicator for male sex, ind_symptom_bowelhabit – indicator for change in bowel habit, MCV_slope_1095 – slope of the mean cell volume (MCV) time series over a 1905 day (3-year) period before FIT, MHC_slope_1095 – slope of the mean cell haemoglobin (MCH) time series, PLT_180 – most recent value of platelets (PLT) in a 180-day window before FIT, ZALB_180 – albumin, CFER_180 – serum ferritin, ind_ZBU_180 – indicator for urea test, PLT_slope_1095 – slope of the platelets time series, MCV_180 – mean cell volume, ind_CB12_180 – indicator for vitamin B12 test, ind_prescription_paracetamol – indicator for paracetamol prescription, MCH_180 – mean cell haemoglobin, MCHC_slope_1095 – slope of the mean cell haemoglobin concentration time series, ATTG_180 – anti-tissue transglutaminase, ZBIL_180 – bilirubin, ZALP_slope_1095 – slope of alkaline phosphatase time series, HGB_slope_1095 – slope of haemoglobin time series.

#### S8. Calibration metrics for ML models

Several calibration statistics—observed/expected ratio, logistic slope (‘calibration slope’), expected calibration error—are reported in Table S8. Of the highest performing models (EBM, NODE-GAM, and NAM), EBM was best calibrated overall with an observed/expected (O/E) ratio of 0.97 (SD = 0.07), logistic slope of 1.05 (SD = 0.09), and expected calibration error (ECE) of 0.09 (SD = 0.04). NODE-GAM tended to under-predict risk with and O/E ratio of 0.88 (SD = 0.19) but had an acceptable calibration slope of 1.02 (SD = 0.07); its ECE was 0.11 (SD = 0.05). NAM under-predicted risk with O/E ratio of 0.94 (SD = 0.26), logistic slope of 1.21 (SD = 0.21), and ECE of 0.14 (SD = 0.06). COLOFIT had an O/E ratio of 1.0 (SD = 0.03), logistic slope of 1.08 (SD = 0.03) and ECE of 0.1 (SD = 0.05).

The ECE measure that was used applied uniform bins over the range of predicted probabilities (e.g. if predicted probabilities ranged from 0 to 0.2, then bins were defined by dividing the [0, 0.2] interval into ten equal-width bins). Further, no weights were applied to bins (bins with more patients were not given more weight). This was important, as the dataset was highly imbalanced (1.2% CRC prevalence), so the majority of predicted probabilities would have been very low and quantile bins would have heavily focussed on the lowest probability region. However, as no weights were applied and some bins may have had a small number of patients, the estimate of ECE may have been noisy.

The cancer rule-in and rule-out performance metrics were the primary focus and these do not require the model be calibrated. However, if any of the models would be further investigated for deployment, calibration could be analysed in more detail, and attempts of recalibration could be made if needed.

**Table S8. Calibration metrics for the machine learning models and COLOFIT**

| Model | Event rate | Mean risk | O/E ratio | Logistic slope | ECE |
|---|---|---|---|---|---|
| COLOFIT | 1.02 | 1.02 (0.03) | 1.0 (0.03) | 1.08 (0.03) | 0.10 (0.05) |
| PLR | 1.02 | 1.12 (0.11) | 0.92 (0.09) | 1.18 (0.18) | 0.09 (0.03) |
| EBM | 1.02 | 1.06 (0.08) | 0.97 (0.07) | 1.05 (0.09) | 0.09 (0.04) |
| NAM | 1.02 | 0.94 (0.26) | 1.16 (0.32) | 1.21 (0.21) | 0.14 (0.06) |
| SNAM | 1.02 | 1.19 (0.37) | 0.94 (0.32) | 1.01 (0.11) | 0.09 (0.07) |
| NODE-GAM | 1.02 | 0.88 (0.19) | 1.22 (0.31) | 1.02 (0.07) | 0.11 (0.05) |
| RF | 1.02 | 1.25 (0.08) | 0.82 (0.05) | 1.09 (0.07) | 0.07 (0.04) |
| XGB | 1.02 | 1.18 (0.54) | 0.98 (0.33) | 1.19 (0.32) | 0.10 (0.04) |
| MLP | 1.02 | 0.91 (0.12) | 1.14 (0.16) | 1.24 (0.41) | 0.08 (0.05) |

O/E ratio - observed/expected ratio; ECE - expected calibration error (computed with uniform bins over the range of predicted probabilities, and no bin-weights).

#### S9. Optimising the models for high areas under the curve

The generalised additive models (GAMs) implemented in pytorch (NAM, SNAM, NODE-GAM) were trained in different ways:

(1) Standard binary cross-entropy (BCE) loss;

(2) Fine-tuning with average-precision (AP) loss after BCE pretraining;

(3) Fine-tuning with AUC-margin loss after BCE pretraining;

(4) Standard BCE loss with an added penalty for hard-positive samples.

The aim was to see whether non-standard strategies (2-4) could produce a better-performing model. These strategies were applied only to GAMs, as these performed the best overall in our analysis (see Results). Due to the software used, the strategies were applicable to GAMs implemented in pytorch.

**AP and AUC-margin losses.** The AP and AUC-margin losses optimise the models for areas under the precision-recall (PR) and ROC-curves, respectively, which may be beneficial in the current dataset with a low prevalence of cancer (approximately 1%). This can be advantageous in low-prevalence datasets, because both areas are maximized when all cancer cases are ranked above all non-cancer cases, whereas the BCE loss can still improve by lowering risk scores of some non-cancer patients even after they are already ranked below every cancer patient. Following the original publications that introduced these losses, the losses were applied after pretraining each model with the BCE loss (see Qi et al, 'Stochastic Optimization of Areas Under Precision-Recall Curves with Provable Convergence' and Yuan et al, 'Large-Scale Robust Deep AUC Maximization', cited in main text).

**Binary cross-entropy with hard-positive penalty.** Since some cancer patients may be harder to distinguish from non-cancers, a penalty term was introduced to encourage the model to focus on these harder cases during training. "Hard positives" were defined as cancer patients whose predicted risk scores fell between the 5th and 20th percentile of all cancer patients' scores (excluding the most extreme cases below the $5^{th}$ percentile while still capturing patients that scored comparatively lower than other cancer patients). The set of hard-positive patients was updated after every training epoch. The hard-positive penalty was formulated as a squared hinge-loss: the mean risk-score of non-cancer patients was subtracted from the mean score of hard positives, and the result subtracted from a margin term (m):

$$L_{hardpos} = \left(m - \left(\mu_{hardpos} - \mu_{neg}\right)\right)^2$$

The overall training loss combined the BCE loss and the penalty term with a tunable weight $\alpha$:

$$\alpha L_{bce} + (1 - \alpha) L_{hardpos}$$

This approach is simple to implement but has a limitation: hard positives are identified using training data, which may fail to capture some genuinely hard cases if the model has overfit.

**Results.** Examining the average precision-recall curves over the cross-validation folds (left column in Figure S9), all training strategies performed similarly. The test-reduction curves were also similar (right column in Figure S8), although for the NAM model, the BCE-pretraining followed by average-precision loss had somewhat lower average reduction in referrals across the sensitivity spectrum and higher variance over the cross-validation folds.

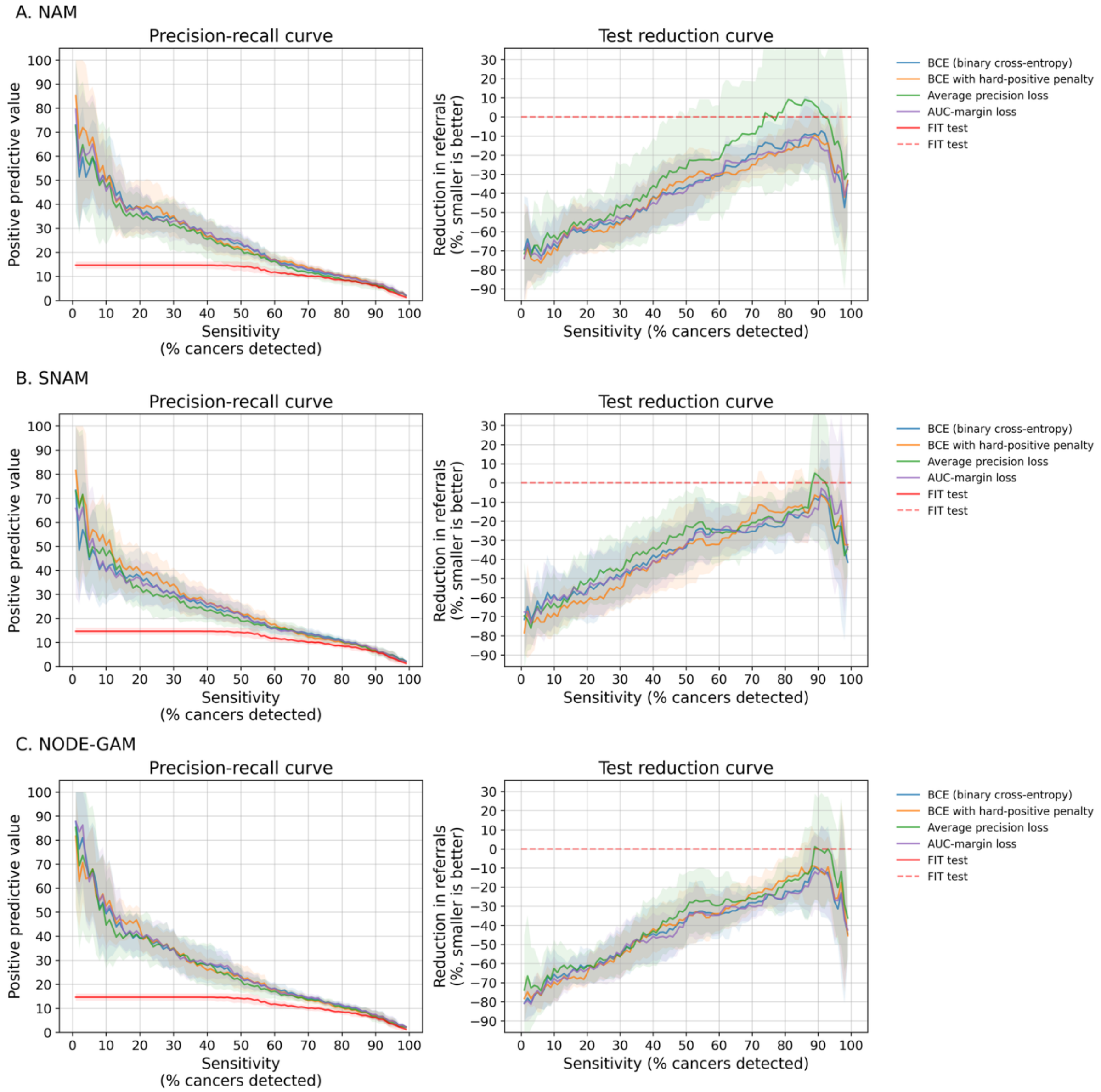

**Figure S9.** Precision-recall curves and test-reduction curves for NAM, SNAM and NODE-GAM models trained using different strategies. BCE: standard binary cross-entropy loss; BCE with hard-positive penalty: BCE loss plus a penalty term for mis-classifying harder-to-detect cancer cases; Average precision loss: BCE pretraining followed by fine-tuning with the average precision loss; AUC-margin loss: BCE pretraining followed by fine-tuning with AUC-margin loss; FIT test: faecal immunochemical test. The test reduction curve approximately shows the percent reduction in referrals (percent reduction in number of patients at or above the risk score threshold) relative to the FIT test when the model is required to have the same sensitivity as the FIT test.

#### S10. Model performance when increasing the number of predictors

Precision-recall (PR) and test reduction (TR) curves for each class of machine learning models, trained with three sets of predictor variables (Sets 1-3), are given in Figure S10. Set 1 contains FIT, age, sex and blood tests available for at least 80% of patients ('common bloods'). Set 2 additionally contains blood test time series slopes for the common bloods over a three-year period before FIT. Set 3 includes additional blood tests and slopes, ethnicity, BMI, diagnosis/procedure/prescription codes (see main text). The NODE-GAM model was only trained on Set 3 to save computational time and is not displayed in Figure S10.

The PR and TR curves of each model trained with three sets of predictors look similar upon visual inspection with some exceptions. For EBM and NAM generalised additive models, the average PPV for Set 3 variables tended to be higher in the 20-50% sensitivity range on the precision-recall curve. For example, for NAM, average PPV at 30% sensitivity was 34.6% (SD = 6.1%) when trained with Set 3 variables, 29.5% (SD = 6.5%) when trained with Set 2, and 27.4% (SD = 7.5%) when trained with Set 1. For EBM, average PPV at 30% sensitivity was 34.6% (SD = 6.5%) for Set 3, 31.7% (SD = 9.5%) for Set 2, and 27.7% (SD = 6.5%) for Set 1. In both EBM and NAM, PPVs at 30% sensitivity were higher under Set 3 compared to Set 1 variables in 7 out of 10 cross-validation folds. Given that the TR curve is computed as (ppv_fit / ppv_model – 1) * 100 at each sensitivity, the reduction in referrals is also higher for Set 3 variables compared to Set 1 variables in the 20-50% sensitivity range for EBM and NAM models.

However, it is not clear whether the differences would remain statistically significant. In this case, statistical testing is not straightforward, because per-fold estimates are not independent, as they were derived from models trained on overlapping sets of data.

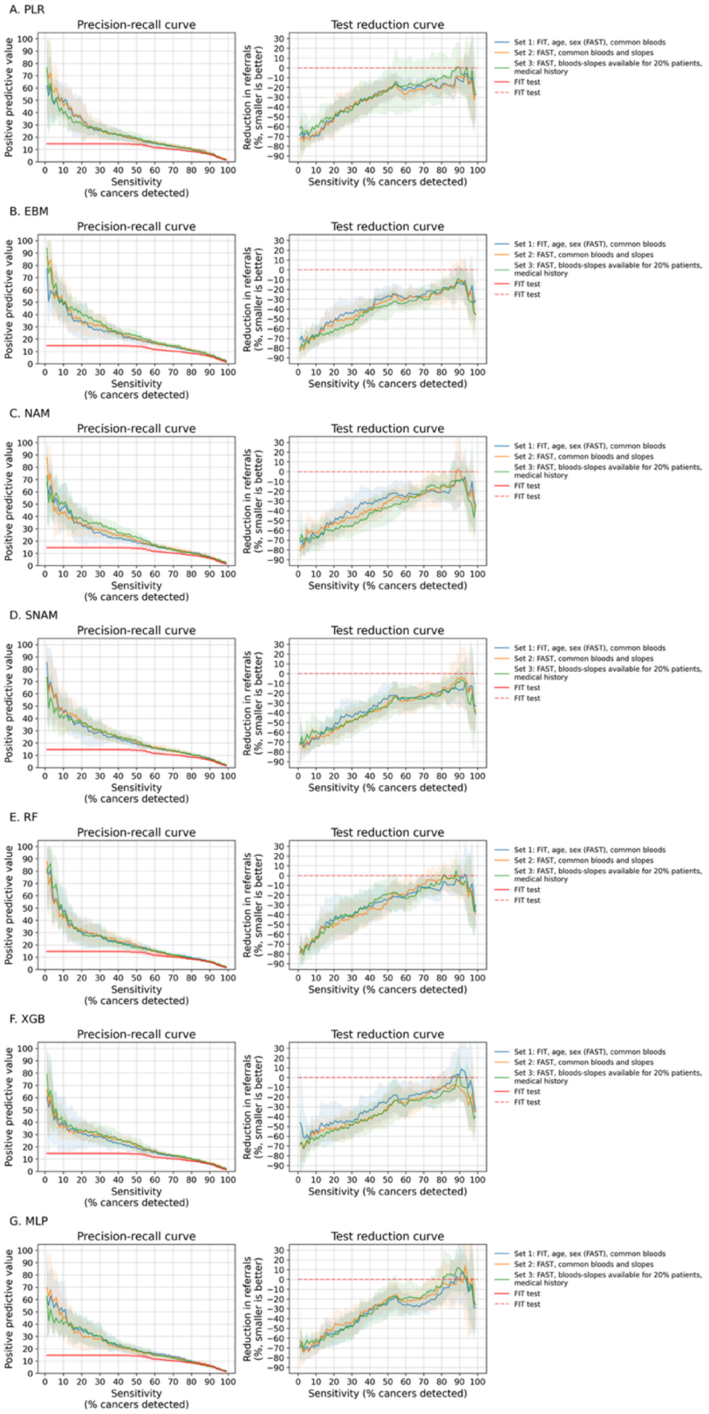

**Figure S10**: Precision-recall and test reduction curves for each machine learning model trained with three sets of predictor variables. The three sets of were (1) FIT, age, sex (FAST) and bloods available for at least 80% of patients ('common bloods'), (2) FAST, common bloods and time-series slopes for common bloods; and (3) set 2, bloods and slopes available for at least 20% of patients; diagnosis/procedure/prescription codes; ethnicity and BMI. Models: PLR – penalised logistic regression, SNAM – sparse neural additive model, NODE-GAM – neural oblivious decision tree ensemble GAM, RF - random forests, GBDT – gradient boosted decision tree, MLP – multilayer perceptron. The curves display the mean (solid line) plus minus standard deviation (shaded area) over the ten cross-validation folds.

#### S11. Sensitivity analysis of the data imputation method

**Motivation.** We conducted a sensitivity analysis to see whether data imputation degraded model performance, focussing on the explainable boosting machine (EBM) generalised additive model (GAM) for two reasons: (1) GAMs performed the best overall (see Results), and (2) EBM can be trained without imputation, by default placing missing values of each variable into a separate 'bin' and estimating their contribution to the linear predictor. Each variable's missing values thus receive their own learned contribution to cancer risk – for example, if patients with missing values have systematically higher or lower cancer risk, the model may be able to learn this. Comparing an EBM trained on imputed data against an EBM trained on data with missing values allowed us to quickly check that the used imputation method (iterative imputing with ridge regression on log-transformed predictors) did not deteriorate model performance compared to a more non-parametric strategy where the contributions of missing values are separately estimated. We explored this on Set 2 predictor variables (FIT, age, sex, common bloods and slopes) and Set 3 predictors (full set) predictor variables. Some bloods and slopes in Set 3 were missing for up to 80% of patients.

**Results.** On Set 2 predictors, EBMs with and without imputation achieved similar average precisions and c-statistics: AP with imputation 25.6% (SD = 3.8%), AP without imputation 26.3% (SD = 3.7%), c-statistic with imputation 94.9% (SD = 1.1%), c-statistic without imputation 95.1% (SD = 1.0%). Reduction in referrals was also similar: 17.5% (SD = 18.2%) with imputation and 18.0% (SD = 14.7%) without imputation. The point estimates in a model without imputation were slightly higher, although the difference is small. On Set 3 predictors, EBMs trained on imputed data and on data with missing values again achieved similar average precisions and c-statistics: AP with imputation 27.7% (SD = 3.0%), AP without imputation 27.8% (SD = 2.7%), c-statistic with imputation 95.0% (SD = 0.7%), c-statistic without imputation 95.1% (SD = 0.7%). Reduction in referrals was higher on average in a model trained on imputed data: 19.7% (SD = 16.4%) with imputation and 11.7% (SD = 18.6%) without imputation.

**Conclusion.** Imputation did not degrade performance in the EBM GAM, and would unlikely to do so in other GAMs (like NODE-GAM), given that GAMs have a similar additive structure.

#### S12. Missingness across the predictor variables

Figure S12 displays the number of observed and missing values for all continuous and ordinal variables in patients with and without colorectal cancer. Categorical variables were handled as follows: sex was not specified for less than ten patients, so an indicator was created for male sex, where 1 indicates “Male” and 0 indicates “Female or not specified” given known higher risk of CRC among males; ethnicity had “Not known” and “Not stated” categories for missing values, which were entered as such into models; diagnosis, procedure, and prescription codes were simply treated as indicators for recorded values.

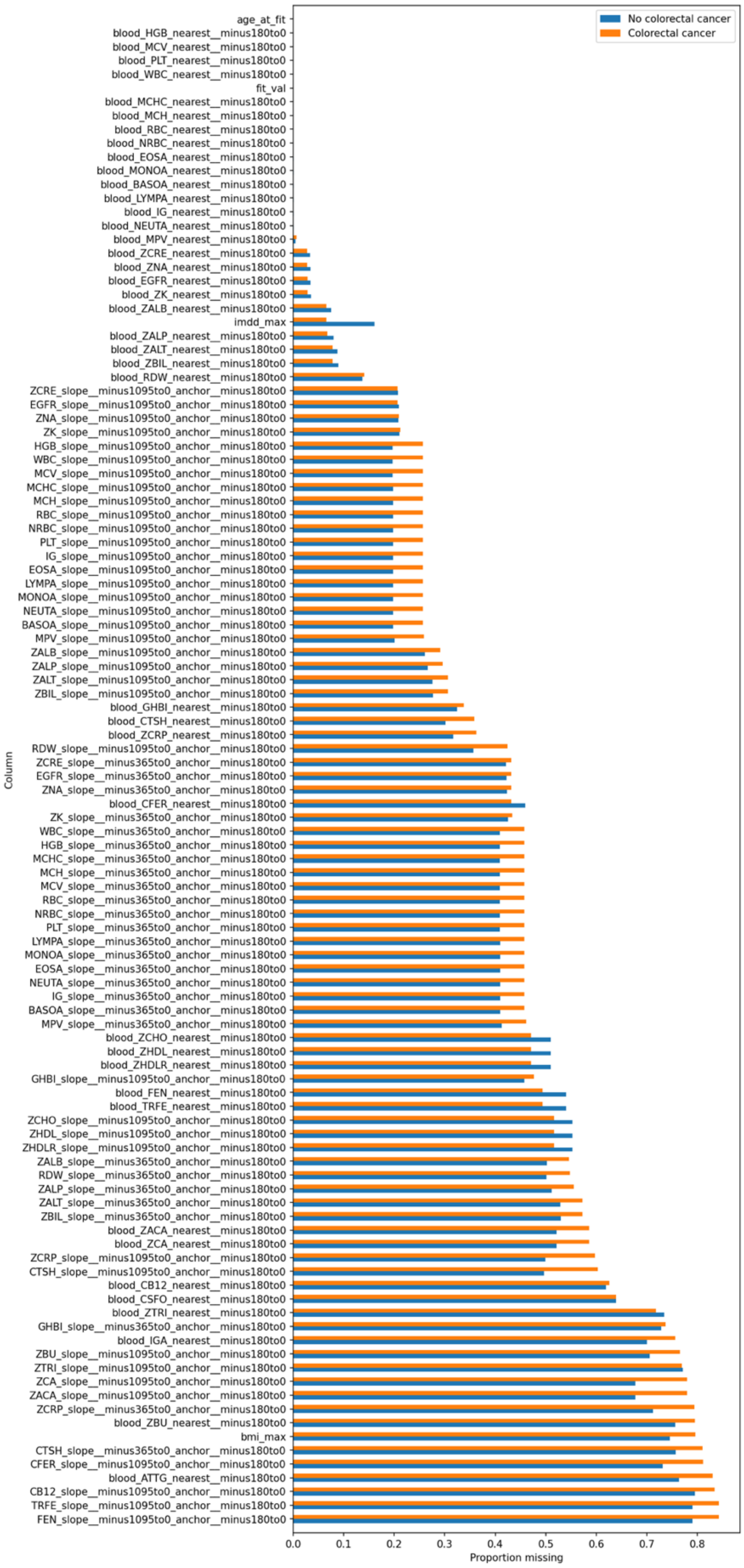

**Figure S12**. Proportion of missing values for continuous and ordinal predictor variables in patients with and without colorectal cancer (CRC). For blood tests, the suffix "_nearest__minus180to0" means "the most recent value within a 180 day time window before the earliest FIT test"; and the suffix "_slope_minus1095to0_anchor__minus180to0" means the slope of the time series within a 1095-day (3 year) time window before the earliest FIT, such that at least one observation occurs within a 180-day window before FIT (the anchor observation). The other time windows for slopes were defined analogously.
